# Antigen receptor stimulation drives selection against pathogenic mtDNA variants that dysregulate lymphocyte responses

**DOI:** 10.1101/2021.10.05.21264464

**Authors:** Jingdian Zhang, Camilla Koolmeister, Jinming Han, Roberta Filograna, Leo Hanke, Monika Àdori, Daniel J. Sheward, Sina Teifel, Yong Liu, Robert A. Harris, Ben Murrell, Gerald Mcinerney, Mike Aoun, Liselotte Bäckdahl, Rikard Holmdahl, Marcin Pekalski, Anna Wedell, Martin Engvall, Anna Wredenberg, Gunilla B. Karlsson Hedestam, Xaquin Castro Dopico, Joanna Rorbach

## Abstract

Pathogenic mitochondrial (mt)DNA molecules can exhibit heteroplasmy in single cells and cause a range of clinical phenotypes, although their contribution to immunity is poorly understood. Here, in mice carrying heteroplasmic C5024T in mt-tRNA^Ala^ – that impairs oxidative phosphorylation – we found a reduced mutation burden in peripheral T and B memory lymphocyte subsets, compared to their naïve counterparts. Furthermore, selection diluting the mutation was induced *in vitro* by triggering T and B cell antigen receptors. While C5024T dysregulated naïve CD8^+^ T cell respiration and metabolic remodeling post-activation, these phenotypes were partially ameliorated by selection. Analogous to mice, peripheral blood memory T and B lymphocyte subsets from human MELAS (Mitochondrial Encephalomyopathy with Lactic Acidosis and Stroke-like episodes) patients – carrying heteroplasmic A3243G in mt-tRNA^Leu^ – displayed a reduced mutation burden, compared to naïve cells. In both humans and mice, mtDNA selection was observed in IgG^+^ antigen-specific B cells after SARS-CoV-2 Spike vaccination, illustrating an on-going process *in vivo*. Taken together, these data illustrate purifying selection of pathogenic mtDNA variants during the oxidative phosphorylation checkpoints of the naïve-memory lymphocyte transition.

**Highlights:**

- In human MELAS patients (A3243G in mt-tRNA^Leu^) and a related mouse model (C5024T in mt-tRNA^Ala^), T and B memory subsets displayed a reduced mtDNA mutation burden compared to their naïve counterparts.
- Selection was observed in antigen-specific IgG^+^ B cells after SARS-CoV-2 Spike protein vaccination.
- T and B cell antigen receptor stimulation triggered purifying selection *in vitro*, facilitating mechanistic studies of mtDNA selection.
- Heteroplasmic pathogenic mutations in mtDNA dysregulated metabolic remodeling after lymphocyte activation and reduced macrophage OXPHOS capacity.

## Introduction

Nuclear and mitochondrial genome (mtDNA) mutations compromising oxidative phosphorylation (OXPHOS) affect tissues with a high aerobic demand, causing multi-systemic and debilitating clinical phenotypes^1–3^. It is well accepted that patients with mitochondrial diseases have an increased infectious disease risk^4,5^, although cellular and molecular studies of the immune system in these individuals are generally lacking.

In recent decades, dedicated efforts have been made to explore the interplay between cellular metabolism and immunology, and mitochondrial metabolism has been found to have critical roles in innate and adaptive immune lineages^6–8^. OXPHOS is essential for powering lymphocyte activation, proliferation and differentiation^9–12^. Naive T lymphocytes fundamentally require elevated OXPHOS and proper mitochondrial reactive oxygen species (mtROS) signaling to undergo antigen receptor activation^10,13,14^. Thereafter, T lymphoblasts interchange between aerobic glycolysis and OXPHOS to fuel rapid proliferation and survival^14^, while substantial spare mitochondrial respiratory capacity is essential for antigen (Ag)-experienced cells to generate long-lived memory cells^7,9,15^, which also use fatty acid oxidation to execute effector functions. Recently, B cells in germinal centers (GCs) that acquired the highest affinity antibody gene mutations were shown to have elevated OXPHOS, and favored fatty acid oxidation over glycolysis^16,17^. Therefore, mitochondrial metabolism is a key requirement for immunological memory. Myeloid cells also depend upon OXHPOS for critical functions, and could also be deleteriously affected by a limited capacity for aerobic respiration^18^, potentially impacting infectious disease susceptibility.

Most immunometabolism studies to date have utilized mitochondrial electron transport chain (ETC) complex inhibitors, or nuclear gene knock-out mice models that completely abolish OXPHOS function in a defined subset of immune cells. Unlike these models, however, maternally-inherited pathogenic mtDNA molecules exhibit heteroplasmy within individual cells and tissues in a process that is poorly understood^19–25^. As each cell harbors many mtDNA copies, when too many pathogenic molecules are present, affected cells can undergo an energy crisis and die as metabolic demands are not met. Importantly, compared to nuclear genome mutations affecting OXPHOS, estimated to be present in 2.9:100,000 individuals, as many as 20:100,000 individuals in populations of European ancestry are predicted to carry pathogenic mtDNA variants^26^. For one of the most common disorders, Mitochondrial Encephalomyopathy with Lactic Acidosis and Stroke-like episodes (MELAS) syndrome, due to the A3243G mutation in *MT-TL1*, prevalence is estimated to be 3.5:100,000^27^. Such mtDNA disorders are incurable and effectively untreatable, showing highly variable penetrance, pathology and prognosis. Thus, animal models carrying heteroplasmic mtDNA mutations are useful translational models for the study of mtDNA-encoded ETC deficiencies. Indeed, pathogenic mtDNA mutations have been targeted for elimination with genetic tools in patient cells and to ameliorate disease phenotypes in C5024T (mt-tRNA^Ala^) mice^28–30^. Despite the clinical importance of such mutations, how such pathogenic polyploidy affects the mammalian immune system is generally unknown, partly because until recently, models such as C5024T mice were unavailable due to technical limitations of engineering the mitochondrial genome.

In this study, we investigated the effects on immune cells of the C5024T mutation in murine tRNA^Ala^, that impairs OXPHOS and recapitulates different aspects of human mitochondrial diseases, such as cardiomyopathy and altered organ mass ratios on the murine C57BL/6N background^29,31,32^. In these animals, mtDNA mutation burden reduction - the percentage by which a tissue had reduced the pathogenic burden compared to baseline – is more pronounced in highly proliferative tissues, such as colonic epithelium and blood^31,33^. A deleterious mtDNA molecule harboring a 3.1-kb deletion in a heteroplasmic *C. elegans* strain^34,35^ also showed increased selection in the intestine, compared to the body wall muscle. It thus follows that different tissues and cell types have variable tolerable mutation thresholds, beyond which a phenotype manifests^31^. Although it is known that proliferation plays an important role in purifying selection, the precise drivers (cues, contexts) and molecular mechanisms of selection in single cells from different tissues remain unknown. Such knowledge in the context of the immune system, could potentially open new avenues for therapeutic targeting of pathogenic mtDNA molecules and processes.

In this respect, we sought to extend our observations in C5024T mice to MELAS syndrome patients who carry different levels of A3243G in another mt-tRNA gene encoding mt-tRNA^Leu(UUR)^. MELAS patients may exhibit severe multi-systemic clinical phenotypes when the heteroplasmy burden is above a given threshold. For example, MELAS patients included in this study – with a baseline mutation burden >90% - suffered from hypertonia, neurological disorders, stroke-like episodes, cardiomyopathy and retinitis pigmentosa, amongst other phenotypes that could have an underlying immunological component^36–38^. In a recent study published during the course of our research, total T and B cells from three MELAS patients were reported to have a lower A3243G mutation burden than in myeloid lineages (which we confirmed), although whether this reduction in highly proliferative immune lineages further delineated between different T and B cell subsets was not addressed^39^. Such a naïve-memory phenotype would have important implications for following the evolution of the peripheral antigen-specific response, or immune ageing^40^, in mitochondrial disease patients, implying the peripheral lymphocyte pool would contain ever fewer fit clones as naïve output declines with age, while memory cell proportions increase^41^.

Based on findings from recent immunometabolism studies, we hypothesized that lymphocytes with high mtDNA mutation burdens are selected against during the transition from naïve to memory cell after antigen encounter, which drives proliferation and elevates aerobic demand, thus shaping the memory antigen-specific repertoire towards low mtDNA burden clones. Using C5024T mice and MELAS patient samples, we demonstrate that T and B lymphocyte populations select against pathogenic mtDNA mutations in response to antigen receptor stimulation both *in vitro* and *in vivo*. This purifying selection illustrates the OXPHOS checkpoints towards immunological memory and reveals molecular phenotypes of heteroplasmic mitochondrial disease.

## Results

C57BL/6 mice with the C5024T mutation (Fig. 1A) and ear heteroplasmy levels between 64-80% at weaning were included in our study. In agreement with previous research showing insidious pathology in this model^31,33^, C5024T mice did not manifest any gross clinical phenotypes with ageing (up to 10-months-of-age in this study), in comparison to wild-type (WT) controls.

**Figure 1.**
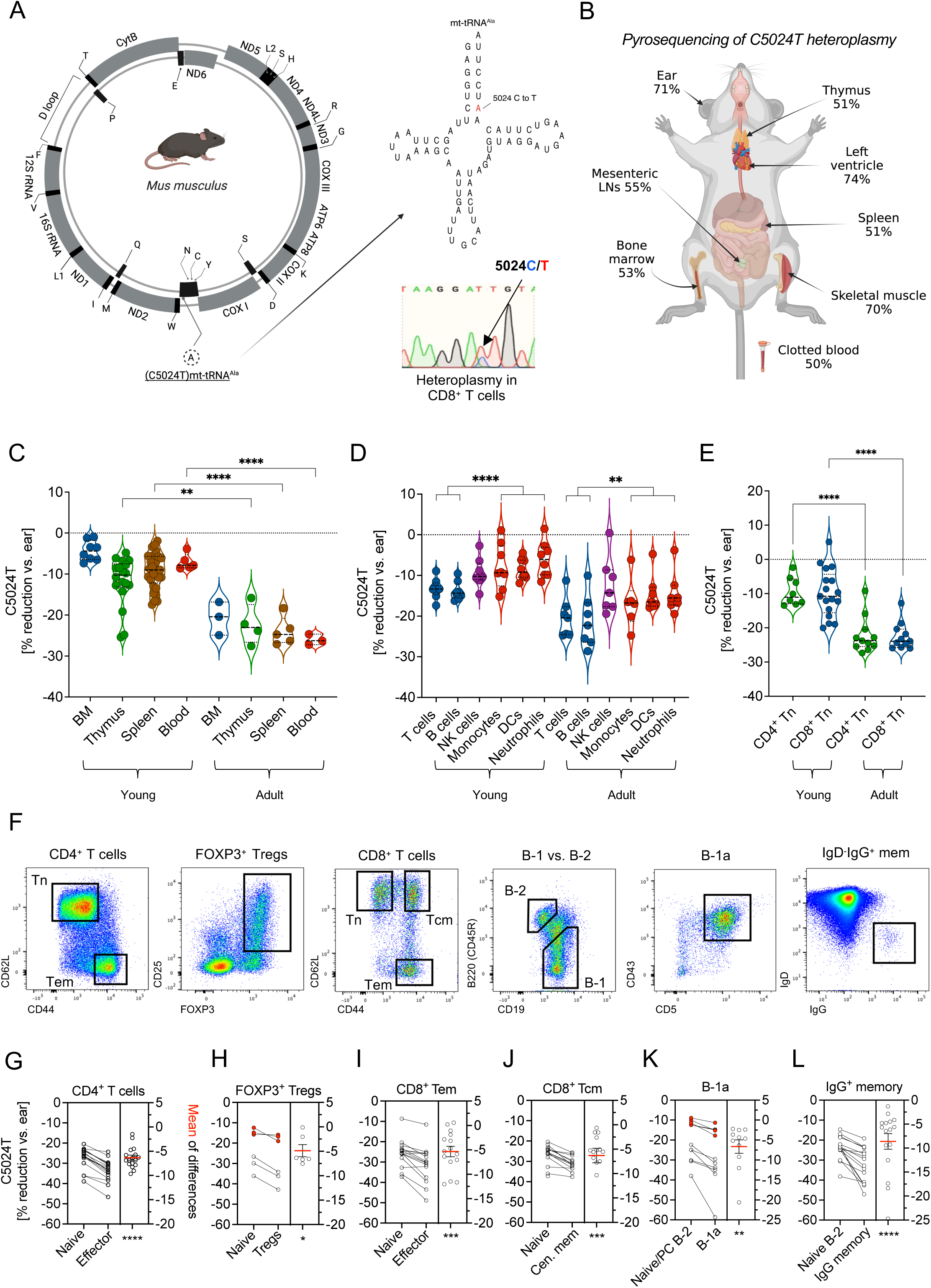
Purifying selection against C5024T in memory T and B lymphocytes. A. Schematic representation of the murine C5024T mutation in mt-tRNA Alanine. An electropherogram showing C/T heteroplasmy in purified naïve CD8^+^ T cells is inset. B. Pyrosequencing results (raw C5024T heteroplasmy percentage in total mtDNA) from different anatomical compartments of a representative, adult (10-month-old) C5024T mouse. *LN: lymph node*. C. C5024T burden reduction – *always the % heteroplasmy reduction relative to ear clip at weaning* - is shown for selected hematopoietic tissues isolated from young (2-month-old) and adult (10-month-old) mice. D. Percentage C5024T burden reduction is shown for selected myeloid (red bars) and lymphoid (blue bars) lineages FACS-isolated from young and adult mice. *DCs: dendritic cells*. E. Percentage C5024T burden reduction is shown for purified naïve CD4^+^ and CD8^+^ T cells from young and adult animals. F. Gates used to FACS-isolate key naïve and memory T and B lymphocyte subsets from C5024T mice, cell surface markers are defined in the text and M&M. *Tn: naïve T cells; Tem: effector memory T cells; Tcm: central memory T cells*. All subsets were isolated from the spleen, apart from B-1a cells, which were isolated from the peritoneum. G. Paired t-test of percentage heteroplasmy reduction for CD4^+^ effector memory vs. naïve in adult mice (*n*=17). The mean of differences in percentage heteroplasmy reduction between both populations is shown for each animal on the right-hand axis. H. Paired t-test of percentage heteroplasmy reduction for CD4^+^ T regulatory vs. naïve; shown in red for three young animals (*n*=6 total). I. Paired t-test of percentage heteroplasmy reduction for CD8^+^ T effector memory vs. naïve in adult mice (*n*=15). J. Paired t-test of percentage heteroplasmy reduction for CD8^+^ T central memory vs. naïve in adult mice (*n*=14). K. Paired t-test of percentage heteroplasmy reduction for B-1a vs. naïve B-2; shown in red for five young animals. *B-1a cells from young animals were compared to peritoneal naïve B-2 cells, otherwise splenic naïve B-2 cells were used* (*n*=11). L. Paired t-test of percentage heteroplasmy reduction for B-2 IgG^+^ memory vs. naïve B-2 (*n*=16).

However, ten-month-old (herein *Adult* mice) but not 2-month-old (herein *Young* mice) male C5024T mice had a ∼15% lower bodyweight than age-matched WT controls (Fig. S1A), as previously reported^33^. A trend of reduced spleen weight was observed for young C5024T mice, although the difference was not statistically significant (Fig. S1A). Furthermore, total white blood cell and erythrocyte counts, and blood Hgb levels, were similar between C5024T and WT animals, as was the cellularity of femur, spleen and thymus at 2-months-of-age (Fig. S1B-C).

Although the thymic distribution of double-positive (DP) and CD4- or CD8-single positive thymocytes was comparable between C5024T and WT animals, the frequency of total CD3^+^ (mature^42^) thymocytes was increased in both young and adult C5024T mice, compared to in controls, suggesting altered thymocyte development in mutant mice (Fig. S1D). Splenic CD4^+^:CD8^+^ T cell ratios, as well as the frequencies of B-2 (CD3^-^CD19^+^B220^+^) and NK (CD3^-^NK1.1^+^) cells, were similar between C5024T mice and WT controls, although the variability increased with age (Fig. S1E). Importantly, we did not observe a significant difference in naïve:effector/central memory ratios amongst splenic T cells in young animals (Fig. S1F), although immunophenotyping of larger sample sizes in adult mice - in consideration of baseline heteroplasmy - is required to address whether the frequencies and absolute numbers of memory cells are altered in this model.

### In vivo selection against C5024T by the innate and adaptive immune systems

C5024T heteroplasmy was determined using a robust mtDNA pyrosequencing assay targeting the murine C5024T mutation that has been extensively validated by us and others^29,31,33^. Briefly, from isolated cell types, PCR was used to amplify a 178-base pair fragment spanning the C5024T point mutation in mtDNA from isolated cell types. PCR products were then purified and denatured to determine the relative levels of mutant and WT mtDNA molecules in a sample using a PyroMark Q24 pyrosequencer (*see Materials and Methods*).

Compared to tissues with a lower proliferative potential in adulthood – such as ear, quadriceps and heart left ventricle – the C5024T burden was more greatly reduced in single cell suspensions from hematopoietic tissues, such as bone marrow, thymus, spleen, mesenteric lymph nodes and peripheral blood (Fig. 1B). Heteroplasmy reduction for each sample of interest was therefore calculated as the percentage change in C5024T frequency, relative to heteroplasmy in the ear (sampled at weaning, *baseline*), as previously described^29^. Previous studies have shown that ear heteroplasmy at the time of weaning is comparable to heteroplasmy in other tissues in aged mice that do not select against the variant^31^.

We found selection against the pathogenic variant across hematopoietic tissues was age-associated, with cells from adult animals having lower mutation burdens than did the same population isolated from young animals (Fig. 1C). These results closely recapitulate mitochondrial disease patients’ heteroplasmy tissue distribution and decreased burden with ageing (for MELAS patients with pathogenic A3243G) ^20,25,43– 46.^

In agreement with a recent study of three MELAS patients (carrying A3243G)^39^, we determined myeloid lineages from C5204T mice to have a higher mutant mtDNA burden than did lymphoid lineages, concurring with their reduced lifespans and proliferation. In C5024T monocytes (CD3^-^CD19^-^CD11b^+^Ly6G^-^Ly6C^hi^), dendritic cells (CD3^-^CD19^-^CD11b^+^CD11c^+^MHC-II^+^) and neutrophils (CD3^-^CD19^-^CD11b^+^Ly6G^+^) sorted from young and adult mice spleens, myeloid selection was also associated with age (Fig 1D and S2A), coinciding with an *in silico* model that predicted selection of A3243G in haemopoietic stem cells^47^. In this respect, age-associated selection of lower mutation burdens was also observed in purified naïve CD4^+^ and CD8^+^ T cells (Fig. 1E).

Together, these data illustrated that circulating lymphocyte precursors and stem cells in the thymus and bone marrow also undergo selection against the mutation, in addition to what might be induced in peripheral immune cells, in agreement with studies showing increased oxidative metabolism in proliferating hematopoietic stem cells (HSCs)^48^, and bursts of high glucose uptake in the common lymphoid progenitor and thymocytes^49^.

### Differential selection of C5024T between naïve and memory T and B cell subsets

As appropriate OXPHOS and mitochondrial functionality is central to lymphocyte activation and the maintenance of T and B cell memory, we hypothesized that mutant mtDNA levels in lymphocytes would delineate by naïve and memory phenotype, with antigen-experienced cells having a reduced pathogenic burden. To test this possibility, we isolated T and B cell subsets of interest by Fluorescence-Activated Cell Sorting (FACS) directly *ex vivo* (Fig. 1F and S2B). As selection became more pronounced with age, and memory cell numbers increased over time, we primarily analyzed the heteroplasmy burden in lymphocytes sorted from adult (10-month-old) animal spleens, with the use of younger (2-month-old) animals being highlighted in the figures. Within the sorted populations from an individual mouse, percentage heteroplasmy reduction for each lineage was calculated relative to ear, as described above. Ear heteroplasmy levels for individual animals are presented in Figure S3.

Within the splenic CD4^+^ T cell compartment of adult mice (*n*=17), we found effector phenotype cells (Tem: CD3^+^CD4^+^CD44^hi^CD62L^lo^) to have a ∼6% lower mutation burden than their naïve (Tn: CD3^+^CD4^+^CD44^lo^CD62L^+^) counterparts (Fig. 1G and S3A), although selection was more pronounced in some animals than others. Furthermore, CD3^+^CD4^+^CD25^+^FOXP3^+^ T-regulatory cells sorted from six animals (*three young and three adult*) had reduced their mutation burden comparably to effector CD4^+^ T cells in adults, in-keeping with their maintenance via autoantigens^50^ and restraining of past effector T cell responses (Fig. 1H and S3B). Indeed, FOXP3^+^ Tregs sorted from these animals were CD44^+^, although cell surface levels were lower than those on effector/memory non-Tregs (Fig. S3B).

A similar pattern was observed in splenic CD8^+^ T cells, with effector (Tem: CD3^+^CD8α^+^CD44^hi^CD62L^lo^) and central memory (Tcm: CD3^+^CD8α^+^CD44^hi^CD62L^hi^) subsets of adult animals (*n=*14) having ∼5% and ∼6% lower mtDNA mutation burdens, respectively, than did naïve (Tn: CD3^+^CD8α^+^CD44^lo^CD62L^hi^) cells (Fig. 1I-J and S3C). Burden reduction was not statistically different between Tem and Tcm subsets.

We next sought to determine whether the same applied to B cells and first isolated fetal-origin B-1a cells (CD3^-^CD19^hi^B220^lo^CD5^+^CD43^+^IgM^+^) from the peritoneal cavity, as these cells – which spontaneously produce natural IgM – are known to depend upon peripheral self-renewal and to have a high capacity for OXPHOS as they protect serosal surfaces^51,52^. Indeed, with found B-1a cells isolated from the peritoneal cavity to have a higher OXPHOS capacity than did naïve B-2 cells (CD3^-^ CD19^+^B220^+^IgM^hi^IgD^hi^) from the same site (Fig. S3D). In agreement with their fetal ontogeny and elevated maximal OCR, B-1a cells had selected more (∼15% reduction) than naïve B-2 cells (∼11% reduction) by 2-months-of-age (Fig. S3E). However, peritoneal naïve B-2 cells selected more than did γδ-T cells (CD3^+^γδ-TCR^+^) isolated from cavity (which reduced mutation burden ∼8% on average), perhaps indicative of the innate-like features of IL-17-producing γδ-T cells that predominate body cavity responses^53,54^ (Fig. S3E). We confirmed these B-1a findings in adult animals, and in one mouse, peritoneal B-1a cells had a striking >20% mutation burden reduction difference to age-selected naïve splenic B-2 cells, which themselves showed a striking 40% burden reduction relative to ear (Fig. 1K).

Amongst splenic B cells, class-switched IgG^+^ B-2 cells (CD3^-^CD19^+^B220^+^IgG^+^IgD^-^ IgM^-^, *n=*16) had a reduced mutation burden compared to un-switched naïve cells (CD3^-^CD19^+^B220^+^IgM^hi^IgD^hi^IgG^-^). (Fig. 1L and S3F). Class-switched IgG^+^ B cells generally showed the strongest naïve vs. memory selection in the subsets studied, averaging an 8.5% difference in burden reduction (Fig. 1L). As observed in T cells, several animals showed more striking (>15%) differences between their naïve and IgG^+^ memory B-2 or B-1a cells, while in others, differences were more subtle, illustrating how selection pressures vary between individuals, perhaps due to environmental effects provoking the immune system.

Taken together, these results demonstrate that the generation/maintenance of immunological memory *in vivo* is associated with purifying selection against a pathogenic mtDNA variant, strongly suggesting the C5024T mutation impairs adaptive immune cell function.

### In vitro stimulation of T and B cells – but not BMDMs – from C5024T mice induces rapid selection

As the T and B memory pools had selected against C5024T *in vivo*, we next sought to determine whether we could induce this process through antigen receptor stimulation *in vitro*, which would represent an OXPHOS checkpoint of immune memory.

To this end, we stimulated naïve CD4^+^ and CD8^+^ T cells via the T cell receptor (TCR) complex (anti-CD3/CD28) and cultured them for 5 days in the presence of IL-2 (Fig. 2A). By staining naïve CD4^+^ and CD8^+^ T cells with a proliferation tracing dye and FACS-isolating cells from different divisions, we observed heteroplasmy burden reduction (purifying selection in the culture) to occur strongly from the third-fourth division onwards (Fig. 2B-C), suggesting that T cells with higher mutation burdens can undergo a few rounds of division under these conditions.

**Figure 2.**
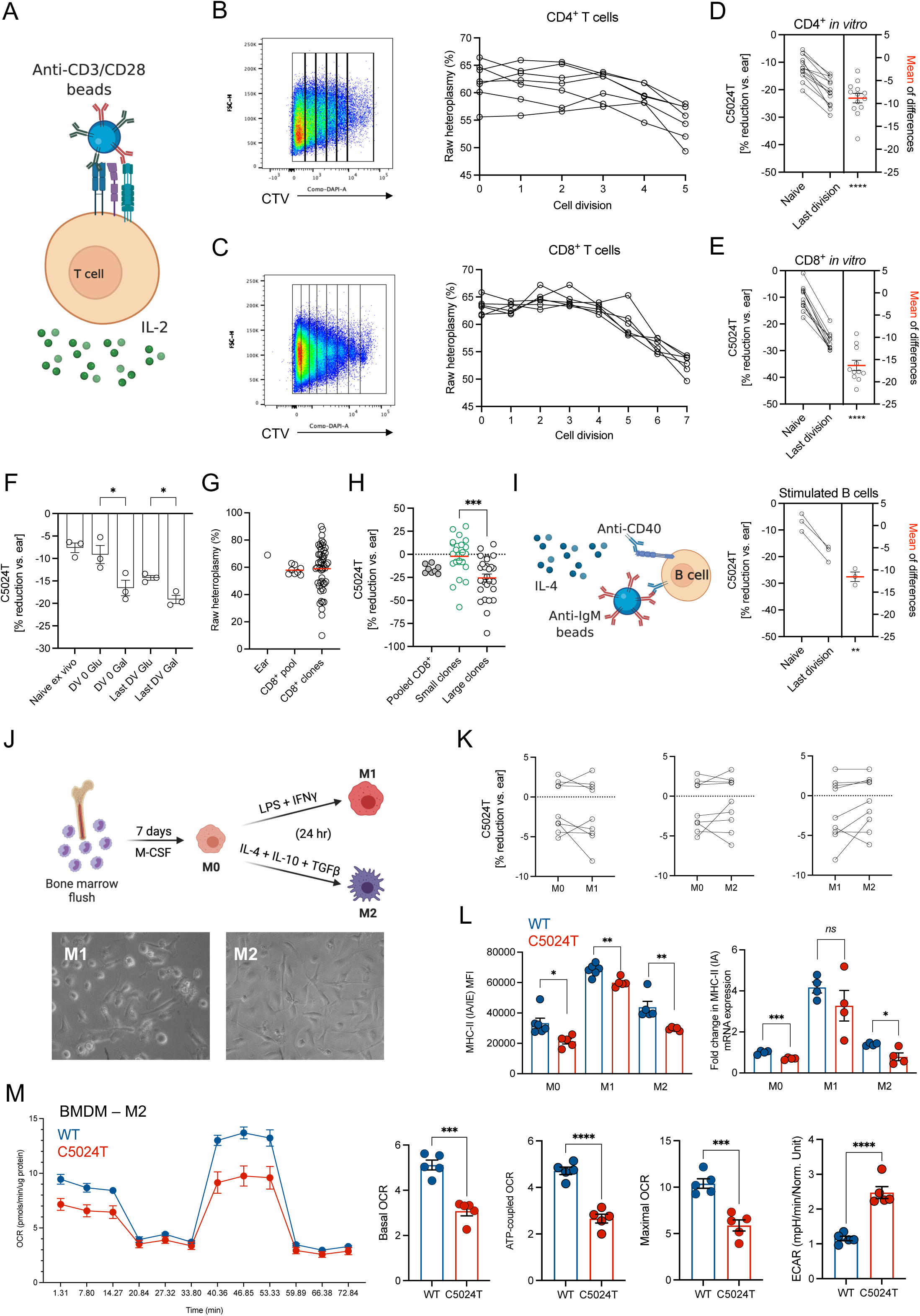
*In vitro* induction of mtDNA selection by antigen-receptor triggering. A. Schematic of T cell stimulation conditions used. Stimulation experiments were carried out in 96-well U-bottom plates with 250,000 cells/well and a 1:1 bead:cell ratio in the presence of 100 U/ml IL-2. B. A cell trace violet (CTV; proliferation dye) pseudocolor dot-plot from *ex vivo*-stimulated naïve CD4^+^ T cells from a representative mouse after 5 days is shown. Raw heteroplasmy frequency is plotted for cells in each division (0-5) sequenced after FACS isolation (*n*=6). C. A cell trace violet (CTV; proliferation dye) pseudocolor dot-plot from *ex vivo* stimulated naïve CD8^+^ T cells from a representative mouse after 5 days is shown. Raw heteroplasmy frequency is plotted for cells in each division (0-7) sequenced after FACS isolation (*n*=6). D. Paired t-test of C5024T heteroplasmy burden reduction in *ex vivo* naïve CD4^+^ T cells vs. stimulated cells sorted from the last division after 5 days stimulation (*n*=13). E. Paired t-test of C5024T heteroplasmy burden reduction in *ex vivo* naïve CD8^+^ T cells vs. stimulated cells sorted from the last division after 5 days stimulation (*n*=11). F. Heteroplasmy burden reduction is shown for purified unstimulated (naïve *ex vivo*) or stimulated naïve CD4^+^ T cells cultured in galactose (Gal) or glucose (Glu) media with IL-2 for 5 days. After 5 days, FACS was used to isolate cells from division (DV) 0 or the last division (*n*=3). G. Raw heteroplasmy percentage distribution in single CD8^+^ T cell clones from a single young mouse, compared to the peripheral CD8^+^ pool and the ear baseline value. *Data are representative of multiple animals* (*n*=5). H. Clonally expanded single CD8^+^ T cells after 7 days of culture post-stimulation were classified as belonging to large (>200 cell/well) or small (<200 cell/well) clonal populations. Total DNA from each well was then subjected to pyrosequencing. The percentage heteroplasmy reduction relative to ear is plotted for each clonal population and the naïve CD8^+^ T cell pool after activation. I. Schematic of conditions used to stimulate murine B cells *in vitro*. A 1:1 bead:cell ratio, 1 ug/ml anti-CD40 and 20ng/ml IL-4 were used in combination for 5 days. Paired t-test of percentage heteroplasmy burden reduction in naïve vs. stimulated cells from the last division (isolated by FACS) is shown in the right-hand panel (*n=*3). J. Schematic of conditions used to polarize macrophages to M0, M1 and M2 states. Images of M1 and M2 cells are shown, confirming typical morphology 24 h after polarization. K. Comparison of heteroplasmy burden change (relative to ear) for M0, M1 or M2 polarized states. No statistically significant changes were observed. L. Flow cytometry staining of MHC class II (clone 2G9 binding I-A^b^, I-A^d^, I-A^q^, I-E^b^, I-E^d^, and I-E^k^) alloantigens on the cell surface of M0, M1 or M2 polarized BMDMs from WT (blue dots, *n*=6) and C5024T mice (red dots, *n*=5). The mean fluorescence intensity (MFI) in the IA/IE channel is plotted (left-hand panel). Fold-change (relative to beta actin) for IA mRNA in M0, M1 and M2 cells from C5024T (*n*=4) and WT (*n*=4) mice (right-hand panel). M. Seahorse Mito Stress Test of C5024T (*n*=5) and WT (*n*=5) M2 BMDM 24 h after polarization. Data normalized according to protein amount/well.

Compared to *ex vivo* naïve CD4^+^ and CD8^+^ T cells collected at day 0, live cells FACS-isolated from the latest division from either lineage (after 5 days of culture) had significantly lower C5024T burdens (Fig. 2D-E). In these cultures, CD8^+^ T cells selected against the variant more strongly than CD4^+^ cells, approximating ∼17% and ∼9%, respectively, in agreement with their increased divisions (Fig. 2B-C) and higher reported dependence on OXPHOS than CD4^+^ cells^9,55^. This sharp decrease in heteroplasmy between *ex vivo* naïve and the most-proliferated CD8^+^ T cells was confirmed by Sanger sequencing (Fig. S4). Selection against the variant was thus more pronounced *in vitro* (∼17% reduction in stimulated CD8^+^ T cells) than was observed *in vivo* (∼5% difference at 10 months-of-age), in agreement with the supraphysiological polyclonal activation conditions of conventional protocols. Furthermore, we found that culturing C5024T CD4^+^ T cells in the presence of galactose (which forces the cells to rely on OXPHOS) exacerbated selection in both un-divided and divided cell populations (Fig. 2F).

To further explore the heteroplasmy-proliferation relationship, we analyzed clonal expansion of C5024T T cells. As reported for other tissues in these mice and mitochondrial disease patients^19,21,23,24,31^, within the peripheral CD8^+^ T cell pool, we found single naïve cells to have a wide range distribution of mutation burden (Fig. 2G). After 8 days of culture-post TCR stimulation, we found larger CD8^+^ clonal populations (>200 cells) to have lower C5024T burdens (∼25% reduction vs. ear) than smaller ones (<200 cells) (∼2% reduction) (Fig. 2H). These results show that even high mutation burden cells – e.g., >75% – can proliferate, although they do so with slower kinetics and relatively little selection. Indeed, compared to WT cells, a greater proportion of live C5024T CD4^+^ and CD8^+^ T cells 5 days post-activation (by which time the population had undergone selection), were in the latest division (Fig. S5A), suggesting cells with higher mutation burdens remain in early divisions or die.

Given our *in vivo* results showing purifying selection in memory B lymphocytes, we also stimulated B-2 cells from the spleens of young mice via the cell surface IgM B cell receptor (BCR) complex in the presence of anti-CD40 and IL-4, and again compared *ex vivo* and most divided states. As in T cells, selection was strongly induced in B cells 5 days after activation and four divisions (Fig. 2I and S5B). The induction of lymphocyte proliferation via the antigen receptor complexes in such models, therefore, facilitates the mechanistic study of mtDNA selection in primary cells.

In contrast to lymphocyte cultures, we did not observe significant differences in pathogenic variant burdens in bone marrow-derived macrophages (BMDMs) polarized to become either M1 (pro-inflammatory; glycolytic, iNOS^hi^CD86^hi^) or M2 (alternatively activating; OXPHOS, Arg1^hi^CD206^hi^)-phenotype cells (Fig. 2J-K and S5C). These results suggested that maturation in the absence of proliferation does not lead to acute selection at the cellular level in BMDMs. This further supported the notion that most of the myeloid burden reduction with age occurs in the self-renewing precursors of circulating cells.

As the degree of heteroplasmy remained similar to ear baseline levels in macrophages polarized under these conditions, it holds that they may suffer functional deficits from the mutation. In this respect, the expression of MHC class II (IA/IE) was found to be significantly lower at the cell surface in C5024T M0, M1 and M2 cells, compared to WT cells (Fig. 2L). This observation with flow cytometry was mirrored at the mRNA level for the class II IA gene but did not reach statistical significance in M1-polarized cells (Fig. 2L). Furthermore, using the Seahorse MitoStress test that characterizes the extracellular metabolic flux, we found C5024T M2 macrophages – which required higher levels of OXPHOS than M1 cells (Fig. S5D) – to have reduced mitochondrial respiration compared to WT cells, and mutant cultures were typified by a high extracellular acidification rate (Fig. 2M and S5E). These results illustrate impaired macrophage function in C5024T mice, which may impact infectious disease susceptibility.

### Dysregulated metabolic remodeling during early T cell activation is partially resolved through purifying selection at the population level

To determine whether the naïve CD8^+^ T cell pool, like M2 macrophages, in C5024T mice had compromised mitochondrial respiration during early activation at the cellular level, we again applied the Seahorse MitoStress test. After 6 hours of CD3/CD28 stimulation (when the heteroplasmy distribution of the naive T cell pool resembled the more variable *ex vivo* state) naïve C5024T CD8^+^ T cells had reduced basal and maximal oxygen consumption rate (OCR) and ATP-coupled OCR (Fig. 3A), compared to WT CD8^+^ T cells, illustrating that the mutation impairs the population response to TCR triggering. However, after 8 days of culture post-TCR activation and a 6-hour re-stimulation with CD3/CD28, these differences were absent, with both C5024T and WT cells responding similarly (Fig. 3B). These results suggest that the T cell population *in vitro* partially restores more optimal OXPHOS through purifying selection after rapid proliferation.

**Figure 3.**
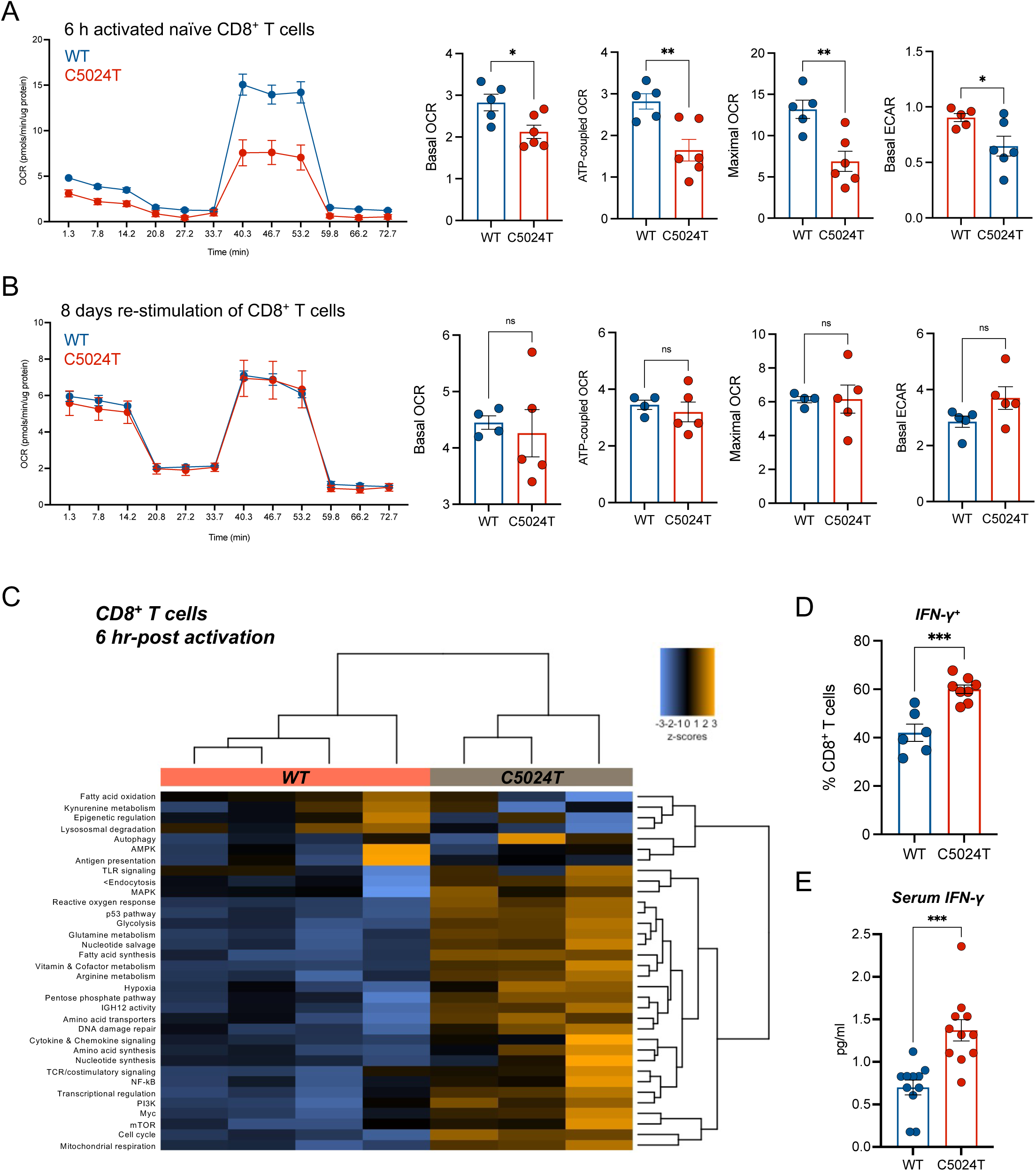
C5024T dysregulates naïve CD8^+^ T cell activation. A. Seahorse Mito Stress Test of *ex vivo* naïve CD8^+^ T cells isolated from young mice (*n*=6 C5024T, *n*=5 WT) and stimulated for 6 h with anti-CD3/CD28 beads and IL-2 before analysis. Data were normalized according to protein amount/well. B. Seahorse Mito Stress Test after re-stimulation CD8^+^ T cells isolated from young mice (*n=5* C5024T, *n*=4 WT). After 8 days of culture with activation at time 0, T cells were counted and re-stimulated for 6 h (anti-CD3/CD28 + IL-2) before Seahorse. C. Nanostring metabolic gene module analysis of mRNA isolated from CD8^+^ T cells from young C5024T (*n*=3) and WT (*n*=4) mice stimulated for 6 h with anti-CD3/CD28 + IL-2. Hierarchical clustering of principle component differences in normalized gene expression per module is shown. D. Proportion of IFN-γ^+^ cells in the latest division, assessed by intracellular flow cytometry after PMA/Ionomycin re-stimulation of CD8^+^ T cells previously expanded for 8 days with anti-CD3/CD28 and IL-2. C5024T (*n*=8) and WT (*n*=6). E. Serum IFN-γ levels in C5024T (*n*=3 adult, *n*=8 young) and WT (*n*=3 adult, *n*=8 young) animals measured by electrochemiluminescence.

To further determine how naïve CD8^+^ T cell cultures were metabolically dysregulated by C5024T, mutant and WT cells were (unstimulated or) stimulated via the TCR for 6 and 96 hours (4 days) and isolated RNA was subjected to transcriptomic analysis using the Nanostring nCounter mouse metabolic pathways panel that analyses the expression of 786 genes across 34 pathways central to cellular metabolism^56^.

Validating our stimulation conditions, within 6 hours of stimulation, the expression of genes important for CD8^+^ T cell effector functions, such as *Il2ra* (CD25), *Tbx21* (T-bet), *Irf4, Gzmb* (Granzyme B) and *Bcl2l1*, were strongly upregulated, compared to time 0 (Fig. S6A). Metabolically and at the same timepoint, the expression of serine hydroxymethyltransferase (*Shmt1*) was strongly induced, confirming a role for one-carbon metabolism in early CD8^+^ T cell activation^57^, while highly-upregulated spermidine synthase (*Srm*) and ornithine decarboxylase (*Odc1*) illustrated the important role recently reported for polyamine metabolism during T cell activation^58^.

Gene module analysis (extracting pathway-level information from a group of genes using the first principal component of their expression data^59^) was used to reveal the overall metabolic remodeling of early T cell activation (Fig. S6B-C). Between the naïve state and 6 hours of activation there was a clear shift from catabolism to anabolism, with decreased AMPK signaling, autophagy and lysosomal degradation, and increased mTOR and Myc signaling. By this time (*6 h*) mitochondrial respiration, aerobic glycolysis, fatty acid synthesis, the pentose phosphate pathway and glutamine metabolism were all sharply upregulated (Fig. S6C). Furthermore, compared to *ex vivo* naïve cells, 6 h-activated CD8^+^ T cells displayed a marked reduction in tryptophan/kynurenine metabolism (Fig. S6C), which can induce apoptosis in T cells^60^. By 96 h post-stimulation, fatty acid synthesis genes were downregulated and networks covering mitochondrial respiration, glycolysis, arginine and glutamine metabolism and fatty acid oxidation were further upregulated, compared to 0 and 6 h, largely consistent with previous research^8,58,61^.

With regards to genotype, in contrast to the *ex vivo* state and the 96 h timepoint, after 6 h of activation C5024T and WT CD8^+^ T cells clustered separately in gene module analysis (Fig. 3C). When genotype was compared at the 6 h timepoint, C5024T cells had a more metabolically active and inflammatory transcriptional phenotype than did WT cells, with increased expression of genes involved in mitochondrial respiration, the cell cycle, cytokine and chemokine signaling, and the ROS and hypoxia responses (Fig. S6D), relative to WT. In contrast, genes involved in epigenetic remodeling were less expressed in C5024T than WT cells. These results highlight a specific pattern of dysregulated metabolic reprogramming of naïve C5024T CD8^+^ T cells after TCR activation. The upregulation of nuclear-encoded metabolic and inflammatory genes (compared to WT cells) perhaps occurs as a compensatory mechanism to improve mitochondrial function in T cells with more severely impaired OXPHOS, as suggested by similar results from fibroblasts from a MELAS (A3243G) patient^62^. Such a compensatory increase could itself dysregulate effector T cell responses.

Consistent with the *in vitro* selection and Seahorse data (Fig. 2B-C and 3A-B), the resolution of the transcriptional clustering by genotype after rapid proliferation (*96 h*) suggested that purifying selection in culture partially relieved some of the genetic dysregulation present in the pool immediately after TCR triggering (Fig. S6B). Nevertheless, to determine whether functional deficits persisted in the CD8^+^ population after proliferation and selection, as suggested by reduced expression of epigenetic remodeling and elevated cytokine signaling genes in C5024T T cells after 6 h activation, we re-stimulated 5 day-activated cultures with PMA/Ionomycin. Six hours after re-stimulation, we found a greater proportion of C5024T CD8^+^ T cells from the latest division to produce IFN-γ, compared to those from WT animals (Fig. 3D), indicating that dysregulated pro-inflammatory responses persist in the T cell population after selection. Furthermore, we found circulating IFN-γ levels to be increased in C5024T mice (Fig. 3E). Increased IFN-γ production has previously been associated with mitochondrial insufficiency and may represent an OXPHOS-glycolysis imbalance in the system that could have pathophysiological consequences^14,63^.

These data indicate that C5024T dysregulates the naïve CD8^+^ T cell response to antigen-receptor stimulation at the transcriptional, protein and cellular levels, and that purifying selection in cultured C5024T cells correlated with partial resolution of phenotypic differences compared to WT cells.

### C5024T mice mount a class-switched neutralizing antibody response after SARS-CoV-2 Spike protein subunit vaccination

To determine whether we could observe a reduction in pathogenic mtDNA burden in response to a specific antigen, and to test whether C5024T mice had an intact T cell-independent B cell response, we inoculated mice with a SARS-CoV-2 Spike (S) glycoprotein trimer vaccine that we have previously studied in mice and other species^64–66^ (Fig. 4A-B).

**Figure 4.**
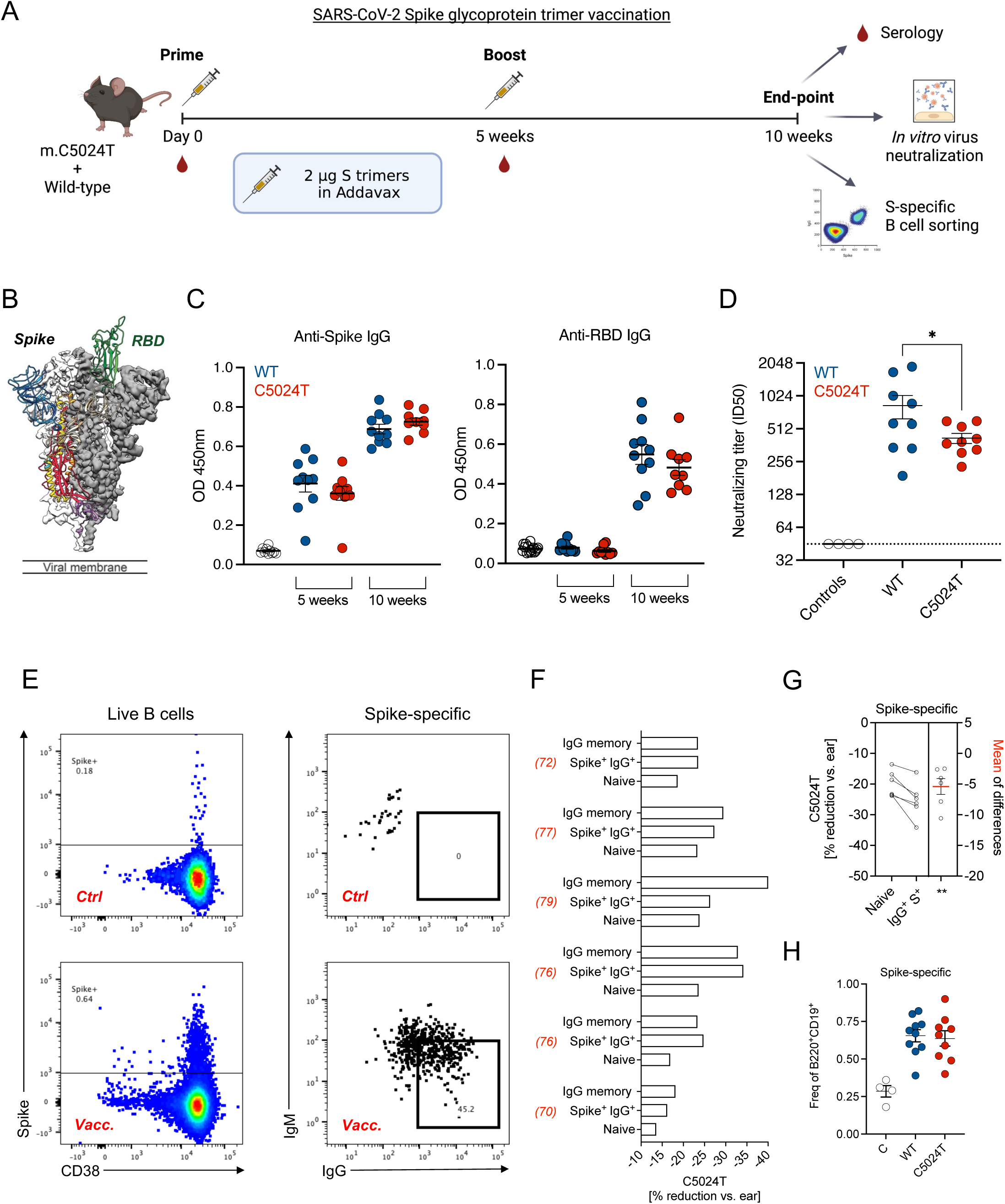
SARS-CoV-2 Spike trimer vaccination of C5024T mice. A. Schematic of the vaccination timeline and readouts. *n=*10 C5024T and *n*=10 age- and sex-matched WT animals (2-months-of-age) were immunized. *One C5024T mouse was sacrificed during the experiment (week 8) due to developing a wound*. B. Cryo-EM structure of the SARS-CoV-2 Spike protein from Wrapp *et al*.^75^, showing the smaller, neutralization-sensitive receptor-binding domain (RBD) in green. C. Anti-S and anti-RBD IgG 5 weeks post-prime (pre-boost) and 5 weeks post-boost. Repeat analysis of unvaccinated controls (*n*=2 WT, *n*=2 C5024T) are shown as open circles. D. ID50 neutralizing titers for vaccinated mice sera. C5024T (*n*=9) shown in red, WT (*n*=9) shown in blue. Unvaccinated controls (*n*=4) are shown at the assay limit of detection. *One WT datapoint was excluded from group analysis for being an outlier (>20-fold above the group mean: ID50 = 20,059)*. E. Flow cytometry strategy for isolation of vaccine-induced class-switched antigen-specific IgG^+^ B cells from the draining lymph node of vaccinated C5024T mice (*n*=6). Spike-binding cells in unvaccinated did not class-switch to IgG, and therefore, likely represent naïve B cells reacting with the probe *in vitro*. Sorting gates are shown in bold outline. F. Heteroplasmy burden reduction (vs. ear) for sorted B cell subsets from vaccinated C5024T mice (*n*=6). Naïve: CD3^-^CD19^+^B220^+^IgD^hi^IgM^hi^IgG^-^; Memory (Ag-specific): CD3^-^CD19^+^B220^+^IgD^-^IgM^-^IgG^+^S^+^; Memory (non-Ag-specific): CD3^-^CD19^+^B220^+^IgD^-^ IgM^-^IgG^+^Spike^-^. Ear heteroplasmy at weaning for each animal is shown in red brackets. G. Paired t-test of percentage heteroplasmy burden reduction for antigen-specific IgG^+^ cells vs. naïve cells. H. Frequency of S-specific B cells from C5024T (*n*=10) and WT (*n*=9) mice plotted as the frequency of Ag-specific from total B-2 cells.

All but one C5024T (from *n*=10) and one WT animal (from *n*=9) developed S-specific IgG responses by five weeks after the priming dose (2 μg S trimers in AddaVax by flank *sub*-*cutaneous* injection), and all had detectable anti-S IgG by five weeks after a homologous boost. IgG specific for the smaller neutralization-sensitive epitope of S, the receptor binding domain (RBD), the predominant target of neutralizing antibodies, was only detectable after the boost dose in all animals (Fig. 4C), and at similar titers in WT and C5024T mice when the AUCs of serial dilutions were analyzed, although these trended towards being reduced in C5024T mice (Fig. 4C and S7A). Despite having similar binding titers at these timepoints, we found that the capacity of vaccinated mice sera to neutralize virus infection *in vitro* was lower for C5024T mice [mean ID_50_ 735 vs. 420 serum dilution] 5 weeks after the boost (Fig. 4D and S7B), suggesting that the quality of the antibody response is negatively affected by the mutation. Thus, although all C5024T mice mounted a neutralizing response after vaccination, further molecular studies are needed to determine whether antibody repertoires are adversely affected by the mutation, e.g., whether vaccine-induced C5024T (vs. WT) antibody lineages show lower levels of somatic hypermutation (SHM) due to a less efficient germinal center response. This might be apparent at the level of virus neutralization, but not at the level of S or RBD binding, as affinity is less of a determinant for the latter. Indeed, no C5024T mice generated ID_50_ neutralizing titers above a 1:600 serum dilution, while 5/10 WT mice did (Fig. 4D).

Post-vaccination differences were also observed in the frequencies of T and B cells between the C5024T and WT mice. C5024T mice had an increased B cell frequency and reduced CD8^+^ T cell frequency in draining (pooled inguinal) lymph nodes 5 weeks after the boost, compared to unvaccinated and vaccinated WT animals (Fig. S7C). However, the frequency of CD4^+^ T cells was not altered. Absolute lymph node cell counts were not significantly different between the mutant and control mice, suggesting that the S vaccine (which *mostly* engaged B cells and not T cells) altered normal lymphocyte proportions *in situ* in C5024T mice.

Our vaccination results, together with our *in vitro* CD8^+^ data, highlight the need to assess vaccine efficacy and infectious disease susceptibility in the C5024T mouse model, despite that this model can mount a neutralizing, class-switched response. We note, however, that in this experiment, young animals (2-month-old) were inoculated^67,68^.

### mtDNA selection in vaccine-induced SARS-CoV-2 S-specific B cells

Five weeks-post boost and using a fluorescently-labelled S probe we generated, we FACS-isolated IgG^+^ S-specific B cells from the draining (inguinal) lymph nodes of vaccinated C5024T mice (*n*=6) to determine their mutation burden (Fig. 4E). Compared to S-negative naïve B-2 cells (CD3^-^CD19^+^B220^+^IgM^hi^IgD^hi^IgG^-^S^-^), class-switched S-reactive cells (CD3^-^CD19^+^B220^+^IgM^hi^IgD^hi^IgG^-^S^+^) had a lower mutation burden (Fig. 4F-G). Thus, these results demonstrate inducible *in vivo* mtDNA selection in peripheral B cells in response to a specific antigen. Finally, the frequency of S-specific cells (from total B cells) in the draining LNs were similar in C5024T and WT mice 5 weeks post-boost (Fig. 4H), in agreement with comparable binding titers.

### Reduction of pathogenic A3243G mtDNA burden in memory T and B cells from human MELAS patients

Given our observations in the heteroplasmic mouse model, we sought to determine whether human memory T and B cells carrying heteroplasmic mtDNA mutations exhibited the same purifying selection observed in mice. For this purpose, we used FACS to examine different peripheral blood immune cell types from two MELAS patients carrying the A3243G mutation in mt-tRNA^Leu^ (Fig. 5A-B and S8-9). Patient clinical characteristics and measured parameters are detailed in Table 1.

**Figure 5.**
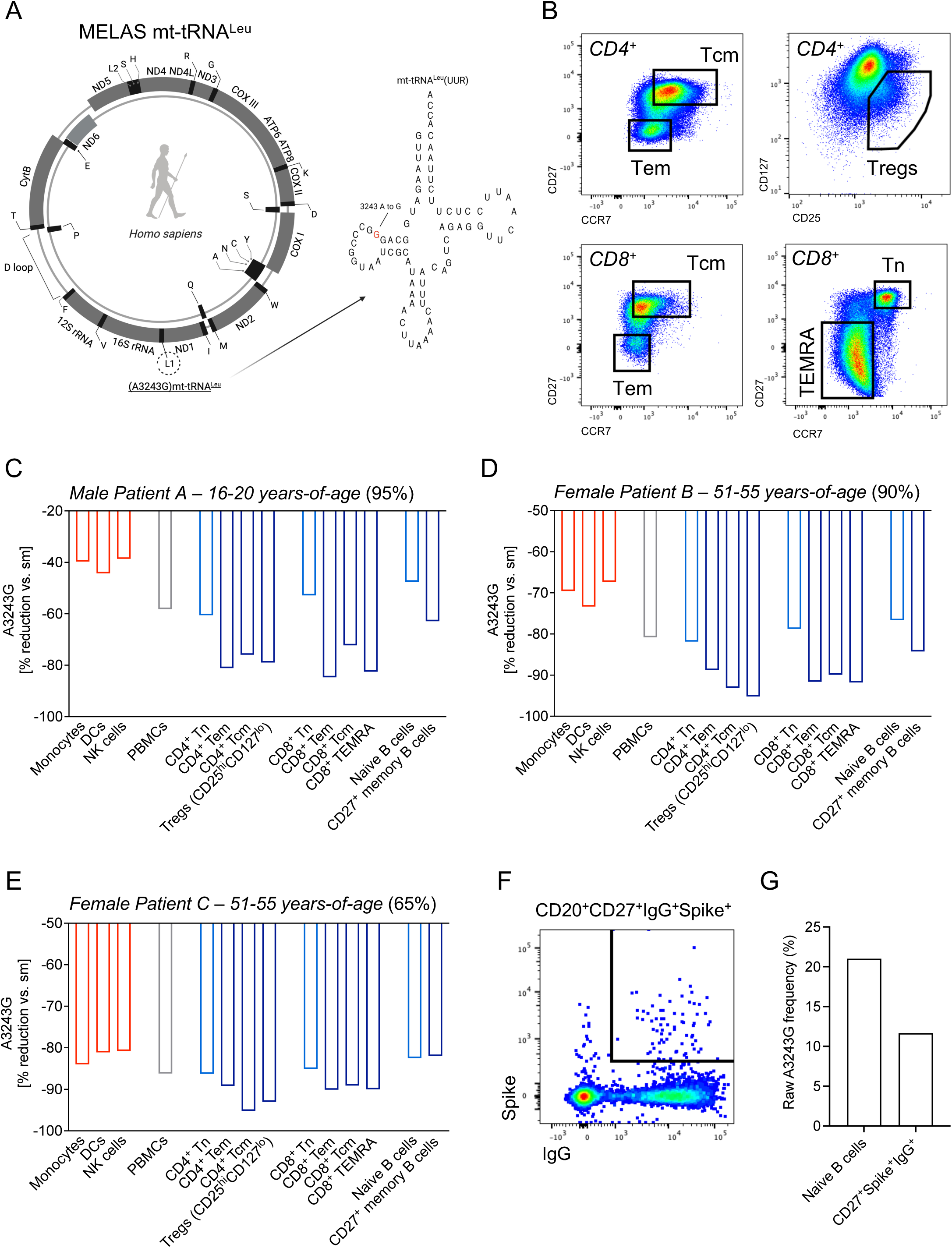
Purifying selection of A3243G in memory T and B lymphocytes from human MELAS patients. A. Schematic representation of the human A3243G mutation in mt-tRNA^Leu^. B. FACS strategy for isolation of key human T lymphocyte populations from MELAS patients. Key T cell subsets are shown, the sorting strategy for all lymphoid and myeloid lineages is shown in the supplementary figures. *Subset markers for FACS are defined in the main text and Materials and Methods*. C. A3243G heteroplasmy burden reduction vs. skeletal muscle (*sm*) is shown for Patient A (male, 16-20 years-of-age). Skeletal muscle heteroplasmy level in patient: 95%. D. A3243G heteroplasmy burden reduction vs. skeletal muscle (*sm*) is shown for Patient B (female, 51-55 years-of-age). Skeletal muscle heteroplasmy level in patient: 90%. E. A3243G heteroplasmy burden reduction vs. skeletal muscle (*sm*) is shown for Patient C (female, 51-55 years-of-age). Skeletal muscle heteroplasmy level in patient: 65%. F. FACS-isolation of antigen-specific memory B cells from Patient B (5 weeks after the first dose of an adenovirus COVID-19 vaccine). Antigen-specific cells were defined as: CD3^-^CD20^+^CD27^+^IgG^+^Spike^+^ and compared to naïve (CD3^-^CD20^+^CD27^-^ IgM^hi^IgG^-^S^-^) cells. The pseudocolor plot of the CD3^-^CD20^+^CD27^+^ B cell population is shown, along with the sorting gate. G. A3243G heteroplasmy frequency (percentage) in naïve and SARS-CoV-2 S-specific B cells from MELAS Patient B.

**Table 1:** MELAS Patients Clinical Information

|  |  |  | **Heteroplasmy level at sampling**<br>(Frequency of A3243G) |  |  |  |
| --- | --- | --- | --- | --- | --- | --- |
|  | **Age** | **Sex** | **Blood** | **Urine** | **Skeletal muscle** | **Clinical phenotype** |
| **Patient A** | 16-20 | Male | 50% | N/A | 95% | Short stature; migraine-like episodes; stroke-like episodes; cardiac arrhythmia; hypertrophic cardiomyopathy; lactic acidosis; neuropsychiatric disorders. |
| **Patient B** | 51-55 | Female | 25% | 80% | 90% | Diabetes mellitus; renal failure; hypertonia; GI motility deficit; migraine-like episodes; hearing deficit; retinitis pigmentosa. |
| **Patient C** | 51-55 | Female | N/A | N/A | 65% | Hearing deficit; exercise intolerance; elevated lactate levels in CSF. |

In MELAS Patient A (male, 16-20 years-of-age), who had a whole blood mutation burden of 50% by age 16-20 (compared to a 95% A3243G burden detected by skeletal muscle biopsy at the same timepoint), we found sorted myeloid lineages to have higher mutant mtDNA burdens than did T and B cells (Fig. 5C), in agreement with a previous study^39^. In this patient, freshly isolated monocytes (CD3^-^CD20^-^CD56^-^ CD14^+^CD16^+^HLA-DR^+^), dendritic cells (CD3^-^CD20^-^CD56^-^CD14^-^CD16^+^HLA-DR^+^) and NK cells (CD3^-^CD20^-^CD56^+^) reduced their A3243G burden by approximately 40%, while total PBMCs reduced their burden by ∼50%.

Within T cell subsets from Patient A, both naïve CD4^+^ and CD8^+^ cells had a higher mutation burden (*∼*60% reduction) than their respective effector, central memory and TEMRA subsets (each approaching an ∼80% reduction) (Fig. 5C). Here, CD8^+^ effector memory cells showed a striking ∼30% greater burden reduction than their naïve counterparts (Fig. 5C). Within the B cells, we also observed naïve (CD3^-^CD20^+^CD27^-^ IgM^hi^) B cells to have a higher mutation burden (−48%) than CD27^+^ memory B cells, with the latter showing a 63% reduction, and were therefore subjected to weaker selection than were memory T cells in this patient.

In non-relative Patient B (female, 51-55 years-of-age with 90% heteroplasmy burden in skeletal muscle, 25% in blood), the same patterns were observed. However, as Patient B was considerably older than Patient A, selection was more pronounced (Fig. 5D). Monocytes, DCs and NK cells from Patient B showed a ∼70% burden reduction at sampling (compared to 40% in Patient A), while PBMCs approximated an 80% reduction.

Despite the strong selection across the hematopoietic system in this patient, naïve T and B cells still had a clearly higher A3243G burden than their memory counterparts. For example, Tregs (CD3^+^CD4^+^CD25^hi^CD127^lo^CD45RA^-^) in this patient showed a striking 95% reduction, while naïve CD4^+^ T cells reduced their burden by 81%. It is noteworthy that mitochondrial complex III is essential for Treg-mediated suppression^69^. CD27^+^ memory B cells also displayed greater selection than did naïve B cells (84 vs. 76% reduction) (Fig. 5D), and as in Patient A, B cells from the female patient also selected against the A3243G variant less strongly than did T cells, perhaps because thymic involution in humans more strongly constricts the peripheral lymphocyte pool than do B cell tolerance mechanisms and output from the bone marrow.

In non-relative female Patient C (female, 51-55 years-of-age), a similar pattern was observed (Fig. 5E). However, this patient had a much lower baseline level of mutation (65% in skeletal muscle) compared to the other patients and differences were more subtle, in line with a less severe clinical history (Table 1). Indeed, the level of mutation in total PBMCs in this patient was 9%, indicating this individual had already undergone strong selection across the hematopoietic system by the time of sampling. As in the other two patients, myeloid lineages selected against the variant less than did total PBMCs, while CD4^+^ and CD8^+^ memory T cell subsets again had a reduced level of mutation compared naïve cells. CD4^+^ T central memory cells in this patient had a heteroplasmy level of just 3%, illustrating very low levels of pathogenic burden in this lineage. We did not, however, observe a difference between naïve and memory B cells in Patient C, perhaps because a ∼10% mutation burden in B cells approaches levels at which the variant is not pathogenic; in agreement with B cells selecting against the variant less strongly than T cells in the patients with higher mutation burdens. Together, these results further highlight the particular need to study the antigen-specific T cell response in MELAS patients.

Finally, at the time of sample collection, five weeks had passed since Patient B had received her first dose of the AstraZeneca COVID-19 adenovirus vaccine. Using ELISA, we found the patient had detectable IgG reactivity to both S and the RBD (Fig. S10), although this was at the mid-low end of the titer range evident in mild (non-hospitalized) adult infections classified using the same assay^70^, in agreement with the patient awaiting her boost dose. Furthermore, using our fluorescent Spike probe, we sorted IgG^+^ S-specific CD27^+^ memory B cells from Patient B, and found them to also exhibit marked selection (∼10% difference in absolute A3243G frequency) with respect to the naïve population (Fig. 5F-G). This illustrates that the selection process was ongoing and was likely induced by vaccination in this case, in agreement with our data from mice. Patient B was not knowingly previously infected with SARS-CoV-2.

As for C5024T, the human A3243G mutation in mt-tRNA^Leu^ is, therefore, also subjected to purifying selection at the naïve-memory lymphocyte transition, further emphasizing the need to study the human and mouse mutations with regards to mitochondrial disease patient immune responses.

## Discussion

As reported by other studies, selection against pathogenic mtDNA variants that dysregulate OXPHOS takes place in hematopoietic stem cells and highly proliferative tissues in an age-dependent manner^31,47^. In addition, selection was observed to take place more-so in lymphocytes than in myeloid cells in samples from three MELAS patients^39^, a phenotype here validated and extended to the C5024T mouse model. These results demonstrate the high metabolic demands of adaptive immunity, which purges the pathogenic mtDNA variants over time, as OXPHOS insufficiency takes its toll on both immature and mature lymphocytes. However, the (i) specific triggers or checkpoints of such purifying selection in stem cells, thymocytes or the peripheral lymphocyte pool; (ii) the relative contributions of these (e.g., central tolerance signaling vs. peripheral antigen encounter); (iii) how these mutations impact on T and B cell responses, have not been previously studied.

Our pyrosequencing data from *ex vivo* mouse and human immune cell subsets revealed that the peripheral lymphocyte naïve-memory transition was accompanied by purifying selection against the pathogenic C5024T and A3243G variants. Therefore, after a lymphocyte has undergone V(D)J recombination and enters circulation, further genetic selection, this time in mtDNA, occurs during the generation/maintenance of adaptive immune memory (and presumably in tandem with somatic hypermutation in B cells). Certain memory T and B cells, like hematopoietic stem cells, are predicted to undergo self-renewal for the lifetime of the organism^71^ and require considerable metabolic remodeling to achieve optimal function and maintain clonal longevity. For example, antigen-experienced B cells are known to require elevated OXPHOS and proliferation to achieve high-affinity antibody maturation in the GC^16^. This may contribute to the slightly reduced neutralizing titers we observed in C5024T mice (vs. WT mice) in response to Spike vaccination, as fewer high affinity anti-RBD clonal lineages with higher levels of somatic hypermutation would be generated, compared to in WT mice, as cells with higher burdens remain in earlier divisions or die as the metabolic demands of the GC response are not met. Thus, the study of vaccine-induced responses and their efficacy, especially taking age into consideration, seems particularly relevant for MELAS and other mitochondrial disorders.

As mtDNA selection in such models progresses with age, presumably until a tolerable level is achieved by a cell lineage and state, it follows that the immune system remains under selective pressure throughout life, especially if the mutation burden is high (e.g., >75% mutation) to begin with. This was evident in the selection observed in class-switched SARS-CoV-2-specific B cells of MELAS Patient B at age 51-55. Selection in the memory cells of Patient B exceeded 90% reductions in certain lineages, compared to skeletal muscle baseline. In humans, the peripheral naïve-memory T cell ratio is approximately equal by the age of 30 and thymic involution makes peripheral self-renewal more important for naïve and memory T lymphocyte maintenance in adulthood and old age^67^. Therefore, in such heteroplasmic models, peripheral self-renewal of ever fewer clonal T and B lineages is predicted to maintain immunity in such patients, as cells with higher mutation burdens fail to fully differentiate and a smaller proportion of the activated pool goes on to form robust memory cells. One consequence could be increased infectious disease susceptibility due to reduced antibody/TCR diversity, especially with ageing, and epidemiological studies of mitochondrial diseases, for long associated with abnormal immune responses^72^, need updating.

With regards to immunological phenotypes associated with the C5024T mutation, our *in vitro* transcriptional and cellular data together indicate that the mutation dysregulates macrophage and T and B cell responses at transcriptional, protein and cellular levels, e.g., by dampening epigenetic regulation of gene expression^58^ and increasing IFN-γ production by activated CD8^+^ T cells^14,63^, while MHC class II cell surface expression and OXPHOS capacity were also significantly reduced in BMDMs. For example, as myeloid cells had a higher tolerance for the mutation, it will be important to decipher how deficits in innate cells contribute to any differences in disease susceptibility in C5024T mice and MELAS patients, e.g., by delaying the initiation of the adaptive immune response.

Importantly, we show that selection is readily inducible *in vitro* by triggering lymphocyte division via the TCR or BCR, providing further evidence that antigenic activation is a major mediator of purifying selection *in vivo*, while facilitating the study of mtDNA selection mechanisms in primary cells. Indeed, as selection was observed in vaccine-induced B cells approximately 5 weeks after immunization, selection takes place shortly after antigen encounter, although homeostatic proliferation and tonic signaling in the periphery likely contribute to ongoing burden reduction in newly formed memory cells over longer periods. Although our studies do not focus on the mechanistic understanding of the selection process itself, several interesting features can be observed that can be important for further analyses. For example, autophagy transcripts were elevated in naïve CD8^+^ T cells and downregulated after TCR triggering (Fig. S6C), while selection occurred strongly from division 4 onwards, suggesting that this pathway is not determinative in the selection process. In addition, we noticed upregulated hypoxia pathway genes in activated C5024T CD8^+^ T cells (compared to WT), while a recent study has reported that oxygen tension contributes to tissue-specific mutation loads^73^. Thus, it is likely that the selection process is influenced by the metabolic environmental conditions in which activated immune cells work, helping to improve their function. It follows that studies of anti-tumor immunity in C5024T mice and mitochondrial disease patients are warranted.

Together, our data reveal an immunophenotype of human mitochondrial disease and support that C5024T mice are a translationally valuable model to study the metabolic requirements of adaptive immunity. Using such models, researchers can further illuminate molecular mtDNA selection mechanisms, energetic bottlenecks in adaptive immune responses, and their clinical consequences.

## Materials and Methods

### Ethical declaration

Ethical approval for the use of peripheral blood samples and clinical information from human MELAS patients was granted by Swedish Ethical Review Authority (registration number 2018/1498-31/3). MELAS patient age boundaries are reported to preserve anonymity. Ethical approval for investigation of C5024T mice, including vaccination challenge, was granted by the Swedish Board of Agriculture (permit numbers 10513-2020, 2001-2018 and 20513-2020). All human and animal studies were carried out in accordance with the guidelines and policies of Karolinska Institutet and EU legislation.

### Animal breeding and housing

C57BL/6NCrl mice harboring a point mutation in pathogenic heteroplasmic C5024T in mt-tRNA^Ala^ were generated as previously described^33^. As C5024T is maternally transmitted, all littermates have different mutation burdens. Control mice (wild-type, WT) with the same nuclear background (C57BL/6NCrl) were used in an age-and sex-matched manner, and animals were housed in a 12-hour light/dark cycle with *ad lib* water and standard chow. Male mice were used throughout, as they were previously reported to have a lower bodyweight than WT mice (http://www.informatics.jax.org/allele/MGI:5902095), whereas females were unaffected in this regard. For *in vivo* and *in vitro* selection experiments, a mixture of male and female mice were used to illustrate selection is sex-independent. Mice were sacrificed using CO_2_ and cervical dislocation at experimental endpoints. Only animals with an ear biopsy heteroplasmy level between 64-80% at weaning were included in this study. Mouse WBC, RBC and total blood hemoglobin parameters were measured using a KX-21N hematology analyzer (Sysmex) according to the manufacturer’s recommendations.

### Human samples

Mitochondrial encephalomyopathy, lactic acidosis, and stroke-like episodes (MELAS) patients under the care of the Center for Congenital Metabolic Diseases (CMMS) at Karolinska University Hospital were invited to participate in this study by their attending clinicians. On the day of clinic visit, peripheral blood samples (20 ml total) were drawn into lithium heparin-coated tubes and processed for PBMC isolation within one hour of venesection. PBMCs were then isolated by density gradient centrifugation over Lymphoprep (StemCell Technologies). Briefly, whole blood was diluted 1:1 in D-PBS with 2% FBS (Cytiva HyClone) and layered onto density gradient medium before centrifugation at 800 *g* for 20 min without brake. An aliquot of plasma was removed prior to gradient centrifugation and stored at -80°C for serological assays before the mononuclear cell layer was collected using Pasteur pipettes. PBMCs were then washed with RPMI-1640 (Cytiva HyClone) with 10% FBS before cryopreservation at - 80°C in FBS with 10% DMSO (Sigma).

### Immunophenotyping and flow cytometry

Primary tissues of interest – spleen, lymph nodes, thymus, femur - were dissected from C5024T or WT mice immediately after sacrifice. To obtain single-cell suspensions, tissues were disrupted/intact femur flushed in D-PBS with 0.2% FBS and filtered through a 70 µm filter before counting with trypan blue using a Countess cell counter (Invitrogen). Body cavity washing was achieved by instilling 8 ml cold D-PBS with 0.2% FBS into the exposed peritoneum before aspiration. For indicated experiments, total B and naïve T cell subsets were purified from splenic single-cell suspensions using magnetic negative selection: B cells were isolated with EasySep Mouse B cell isolation kit (StemCell Technologies); T cells were isolated using naïve CD4^+^ or CD8a^+^ T cell isolation kits (Miltenyi Biotech.). Otherwise, single-cell suspensions were re-suspended to the appropriate concentration and stained with fluorescently labelled monoclonal antibodies in the dark for 45 min at 4°C (please see Results and Resource Table). For intracellular protein staining, samples were permeabilized, fixed and stained using the Foxp3/Transcription Factor Staining Buffer Set (Invitrogen) according to the manufacturer’s instructions. Aqua or near-IR live/dead dyes (Invitrogen) were used to discriminate viable cells by flow/FACS, while CFSE (eBioscience) or Cell Trace Violet (ThermoFisher Scientific) dyes were used to assess proliferation, according to the manufacturer’s instructions. Cells were fixed for analysis (but not FACS) using 4% PFA.

Flow cytometry panels were combinations of the monoclonals listed in the Supplementary Methods. Live-discriminated (splenic) murine cells were defined as follows:

Monocytes: CD3^-^CD19^-^CD11b^+^Ly6G^-^Ly6C^hi^
Neutrophils: CD3^-^CD19^-^CD11b^+^Ly6G^+^
Dendritic cells: CD3^-^CD19^-^CD11b^+^CD11c^+^MHC-II^+^
NK cells: CD3^-^CD19^-^NK1.1^+^
CD4^+^ T naïve: CD3^+^CD4^+^CD44^-^CD62L^+^
CD4^+^ T effector: CD3^+^CD4^+^CD44^hi^CD62L^-^
CD4^+^ Tregs: CD3^+^CD4^+^CD44^+^CD25^+^FOXP3^+^
CD8^+^ T naïve: CD3^+^CD8^+^CD44^-^CD62L^+^
CD8^+^ T effector: CD3^+^CD8^+^CD44^hi^CD62L^-^
CD8^+^ T central memory: CD3^+^CD8^+^CD44^hi^CD62L^+^
γδ-T cells: CD19^-^CD3^+^γδ-TCR^+^
B-1a: CD19^hi^B220^lo^CD5^+^CD43^+^IgM^+^
Naïve B-2: CD3^-^CD19^+^B220^+^IgD^hi^IgM^hi^IgG^-^
Memory B-2: CD3^-^CD19^+^B220^+^IgD^-^IgM^-^IgG^+^
Antigen-specific B-2: CD3^-^CD19^+^B220^+^IgM^-^IgG^+^Spike^+^

A mAb cocktail of FITC-conjugated anti-mouse IgG1, 2b and 3 subclasses was used to identify class-switched B cells. The in-house Spike probe was used at a 1:1000 dilution alongside other cell-surface antigens.

Flow cytometry data were acquired on BD FACSCelesta and BD FACSAria Fusion instruments (BD Biosciences), or an Attune Acoustic Focusing Cytometer (Invitrogen). FACS was carried out using a 16-colour FACSAria Fusion. A minimum of 150 cells were used to determine heteroplasmy in antigen-specific cells, while >2,500 cells were sorted for all other human/mouse subsets of interest. Flow/FACS data were analyzed using FlowJo v.10 software (TreeStar Inc.).

Serum IFN-γ was measured according to the manufacturer’s recommendation using the MSD electrochemiluminescence platform (MesoScale Discovery).

### MELAS patient T and B cell sorting

After thawing frozen PBMCs aliquots, cells were washed with RPMI complemented with 10% FBS before resting at 37°C (5% CO_2_) for 30 min. Cells were pre-stained for CCR7 during incubation, as recommended for this antigen; all additional staining was carried out at 4°C after PBMC resting post-thaw. Cells were stained with Live/Dead dye for 15 min prior to surface staining.

The following panels were used to classify human cells:

Monocytes: CD45^+^CD3^-^CD20^-^CD56^-^CD14^+^CD16^+^HLA-DR^+^
Dendritic cells: CD45^+^CD3^-^CD20^-^CD56^-^CD14^-^CD16^+^HLA-DR^+^
NK cells: CD45^+^CD3^-^CD20^-^CD56^+^
CD4^+^ T naïve: CD45^+^CD3^+^CD20^-^CD4^+^CD45RA^+^CCR7^+^
CD4^+^ T effector memory: CD45^+^CD3^+^CD20^-^CD4^+^CD45RA^-^CD27^low^CCR7^low^
CD4^+^ T central memory: CD45^+^CD3^+^CD20^-^CD4^+^CD45RA^-^CD27^hi^CCR7^hi^
CD4^+^ Tregs: CD45^+^CD3^+^CD20^-^CD4^+^CD45RA^-^CD127^low^CD25^+^
CD8^+^ T naïve: CD45^+^CD3^+^CD20^-^CD8^+^CD45RA^+^CCR7^+^
CD8^+^ T effector memory:CD45^+^CD3^+^CD20^-^CD8^+^CD45RA^-^CD27^low^CCR7^low^
CD8^+^ T central memory: CD45^+^CD3^+^CD20^-^CD8^+^CD45RA^-^CD27^hi^CCR7^hi^
CD8^+^ TEMRA: CD45^+^CD3^+^CD20^-^CD8^+^CD45RA^+^CD27^low^CCR7^low^
Naïve B cells: CD45^+^CD3-CD20+CD27^-^IgM^hi^IgG^-^
Memory B cells: CD45^+^CD3-CD20+CD27+IgM^low^
SARS-CoV-2 Spike-specific B cells: CD45^+^CD3^-^CD20^+^CD27^+^IgM^-^IgG^+^Spike^+^

### In vitro stimulation of mouse T and B cells

Naïve CD4^+^ or CD8^+^ T cells and resting B cells were enriched from splenocytes isolated from 2-month-old mice, as described above. Isolated naïve T cells were stained with CFSE and 250,000 cells per well were seeded on 96-well U-bottom plates with/without a 1:1 (cell:bead) ratio of Mouse T-Activator CD3/CD28 beads (Invitrogen) in complete RPMI-1640 (with 10% FBS, 2.05 mM L-glutamine (Sigma), 1% penicillin/streptomycin (Sigma), 100 U/ml IL-2 (Peprotech) and 55 μM 2-ME (Gibco)). For some experiments, naïve CD4^+^ T cells were also stimulated in complete RPMI-1640 media with no glucose supplemented with 10 mM galactose. Polyclonal T cell stimulation of whole spleen suspensions was done using plate-bound anti-CD3 (5 μg/ml, clone 145-2C11, BD Biosciences) and anti-CD28 (2 μg/ml, clone 37.51, Biolegend) in 24-well plates at a density of 2×10^6^ cells/well. In re-stimulation experiments, PMA/Ionomycin was added for 1 h before protein transport was inhibited for 4 h (1X Cell stimulation/Inhibitor Cocktail, eBioScience). Samples were then washed and stained for intracellular antigens before analysis by flow cytometry.

For selected clonal expansion experiments, modified from Lemaître *et al*^74^, purified naïve CD8^+^ T cells from young C5024T mice were first activated using 1:1 ratio of anti-CD3/28 beads in the presence of 100 U/ml IL2. After 24 h activation in the pool, CD8^+^ T cells were plated (in the presence of IL-2) by limiting dilution (0.8 cell/well) into 96-well (U-bottom) plates pre-coated with anti-CD3/CD28. After limiting dilution, each well was examined under microscope and only wells containing a single cell were marked for further analysis. The four corner-wells of each plate were seeded with 5,000 cells, serving as pooled activation wells. After 7 days clonal expansion, marked wells were imaged for cell counting and lysed, before DNA extraction and pyrosequencing.

250,000 purified resting B cells were labelled with CSFE before activation using a 1:1 (cell:bead) ratio Dynabeads Rat anti-Mouse IgM (Invitrogen), anti-CD40 (HM40-3, 1 μg/ml, BD Biosciences) and IL-4 (20 ng/ml, R&D Systems) in complete RPMI-1640 culture media.

### Bone marrow-derived macrophage (BMDM) in vitro differentiation & polarization

Femurs were dissected intact from recently terminated animals and kept in ice-cold DMEM (Cytiva HyClone) with 0.2% FBS. Bone-marrow cells were collected by cutting off the head of the femur at the neck and flushing the cavity using 27-gauge needle, before filtering through a 70 μm strainer. Cells were washed and centrifuged before being re-suspended in DMEM with 20% FCS, 100 U/ml penicillin/ streptomycin, 2 mM L-glutamine, 20 μM 2-ME, 20 ng/ml recombinant mouse M-CSF in T75 flasks and incubated. Media was changed at days 4 and 7. Mature BMDM were harvested using pre-warmed EDTA at a concentration of 2 mM (30-40 min, 37°C) and evaluated by flow cytometry for expression of CD11b and F4/80. Confirmed mature BMDM cells were then seeded in 6-well plates and after attachment, M1/M2 stimulation medium was added for another 24 h. M1 polarization: DMEM with 10% FBS, 50 ng/ml LPS (O55:B5, Sigma), 20 ng/ml IFN-γ. M2 polarization: DMEM with 10% FBS, 20 ng/ml IL-4, 20 ng/ml IL-10, 20 ng/ml TGF-β. All recombinant mouse cytokines were purchased from Peprotech.

### Extracellular Metabolic Flux Assay

The Seahorse XFe96 Analyzer (Seahorse Bioscience) was used to measure oxygen consumption rate (OCR) and extracellular acidification rate (ECAR) in B-1a and B-2 cells, CD8^+^ T cells and macrophages. For non-adhesive lymphocytes, cells were washed with Seahorse XF RPMI assay medium, then 200,000 cells per well were seeded (in triplicate) in 40 μl assay medium in XF 96-well cell culture microplate coated with poly-D-lysine (Sigma). The plate was centrifuged at 300 *g* for 5 sec with no brake, rotated 180° and centrifuged again for 5 sec at 300 *g*. After centrifugation, 140 μl of assay medium was added per well and the plate was left to stabilize in a 37°C, non-CO_2_ incubator for 40 min. For BMDMs, 30,000 cells were seeded, cultured for 24 h with IFN-γ, LPS or IL4 to polarize them into M1 or M2 subsets. During seahorse, wells were sequentially injected with compounds to achieve final concentrations of 1,264 μM oligomycin (Sigma); 2μM FCCP (Sigma); 0.5 μM rotenone (Sigma) together with 0.5 μM antimycin A (Sigma). OCR and ECAR were measured for each well three times, every three minutes, before and after each injection. OCR was normalized to protein concentration using a BCA Protein Assay kit (ThermoFisher Scientific) conducted according to the manufacturer’s instructions.

### Quantification of mtDNA point mutation heteroplasmy levels

Heteroplasmy levels of C5024T (mouse) or A3243G (human) mtDNA point mutations were determined by pyrosequencing, as previously described^29^. Briefly, for the C5024T mutation, a 178–base pair mtDNA fragment spanning the C5024T point mutation was PCR-amplified using a biotinylated forward primer (5′ TTCCACCCTAGCTATCATAAGC) and a non-biotinylated reverse primer (5′ GTAGGTTTAATTCCTGCCAATCT). For the human mutation the following primers were used: forward, non-biotinylated 5′ CCTCCCTGTACGAAAGGACA; reverse, biotinylated 5′ TGGCCATGGGTATGTTGTTA. After adding Streptavidin Sepharose TM High-Performance beads (GE Healthcare), PCR products were purified and denatured using a PyroMark Q24 vacuum workstation (Qiagen). Sequencing was carried out with PyroMark Gold Q24 reagents and sequencer (Qiagen) according to manufacturer’s recommendations and using the sequencing primers 5′ TGTAGGATGAAGTCTTACA (for mouse) and 5′ GGTTTGTTAAGATGGCAG (for human).

### SARS-CoV-2 immunogen and probe generation

SARS-CoV-2 spike trimers stabilized in the prefusion conformation with either 2 prolines^75^ or 6 prolines^76^ were produced as in Hanke *et al*^65^. Briefly, the plasmids were used to transiently transfect FreeStyle 293F cells using FreeStyle MAX reagent (ThermoFisher Scientific). The spike was purified from filtered supernatant on Streptactin XT resin (IBA Lifesciences) or His-Pur Ni-NTA resin (ThermoFisher Scientific), followed by size-exclusion chromatography on a Superdex 200. The RBD domain (RVQ–VNF) of the emergent strain^77^ spike protein was cloned upstream of a sortase A recognition site (LPETG) and a 6xHis tag and expressed in 293F cells, as described above. RBD-His was purified from filtered supernatant on His-Pur Ni-NTA resin (ThermoFisher Scientific), followed by size-exclusion chromatography on a Superdex 200. Fluorescent spike was generated by first attaching dibenzocyclooctyine-*N*-hydroxysuccinimidyl ester (DBCO-NHS, Sigma) to the spike trimer in a 3:1 molar ratio before attaching AbberiorStar-635P-azide (Abberior GMBH) by strain-promoted azide-alkyne cycloaddition. The final product was purified from unreacted DBCO and fluorophore using a PD-10 desalting column (Cytiva HyClone).

### SARS-CoV-2 spike vaccination

Ten WT and 10 C5024T male animals of 2-months-of-age were used; one C5024T animal had to be sacrificed before the end of the experiment (week 8) because of infection. Immunogens were aliquoted and diluted immediately before administration. Animals were injected *sub-cutaneously* over the inguinal lymph nodes with 100 μl of 2 μg Spike trimers in PBS plus AddaVax (Invivogen) at day 0 (prime dose) and after 5 weeks (boost dose). Two WT and two C5024T mice were injected with sterile PBS for use as un-vaccinated controls. Tail vein blood was collected 5 weeks after the prime dose (pre-boost) and 5 weeks after the boost dose (at week 10, endpoint). After blood clotting at room temperature, sera were separated by centrifugation (10 min, 6,000 *g*) and stored at −80°C until use. Sera were heat-inactivated for virus neutralization experiments.

### Anti-SARS-CoV-2 ELISA

Mice sera collected from different timepoints were tested for Spike- and RBD-specific IgG by ELISA, as previously described^64^. Briefly, 96-well ELISA plates (Nunc MaxiSorp, ThermoFisher Scientific) were coated with freshly prepared SARS-CoV-2 S trimers or RBD (100 µl of 1 ng/µl) in PBS for 15 h at 4°C. Plates were washed six times with PBS–Tween-20 (0.05%) and blocked using PBS-5% non-fat milk (blocking buffer, Sigma). Mice serum samples were thawed at room temperature, diluted, vortexed and incubated in blocking buffer for 1 h (4°C) before plating to block non-specific binding. Serum samples were incubated for 15 h at 4°C to allow low-affinity binding interactions, before washing as before. HRP-conjugated goat anti-mouse IgG (clone 1036-05, Southern Biotech) was diluted 1:2,000 in blocking buffer and incubated with samples for 1 hour at 4°C. Plates were washed a final time before development with TMB Stabilized Chromogen kept at 4°C (Invitrogen). The reaction was stopped using 1 M sulphuric acid and optical density (OD) values were measured at 450 nm using an Asys Expert 96 ELISA reader (Biochrom Ltd.). Secondary goat anti-mouse IgG (Southern Biotech, clone 1036-05) was used at 1:2,000 dilution. All assays were developed for their fixed time, and negative control samples (unvaccinated mice sera) were run alongside test samples. Anti-SARS-CoV-2 S and RBD IgG were detectable at up to 1:10,000 serum dilution using this assay. All samples were run at 1:100 serum dilution, unless otherwise indicated. Human MELAS Patient B anti-S and -RBD IgG reactivity was analyzed by ELISA as we have previously described^70^.

### In vitro virus neutralization assay

Pseudotyped SARS-CoV-2 viruses were generated by the co-transfection of HEK293T cells with plasmids encoding the SARS-CoV-2 spike protein harboring an 18 amino acid truncation of the cytoplasmic tail; a transfer plasmid encoding firefly luciferase; and a lentiviral packaging plasmid (Addgene) using Lipofectamine 3000 (Invitrogen). Media was changed 12–16 h post-transfection and pseudotyped viruses harvested at 48 and 72 h, clarified by centrifugation and stored at −80°C until use. Pseudotyped neutralization assays were adapted from protocols validated to characterize the neutralization of HIV, but with the use of ACE2-expressing HEK293T cells. Briefly, pseudotyped viruses sufficient to generate ∼100,000 RLUs were incubated with serial dilutions of heat-inactivated serum for 60 min at 37°C. Approximately 15,000 HEK293T-ACE2 cells were then added to each well and the plates incubated at 37°C for 48 h. Luminescence was measured using Bright-Glo (Promega) according to the manufacturer’s instructions on a GM-2000 luminometer (Promega) with an integration time of 0.3s. The neutralization assay limit of detection was at 1:45 serum dilution and ID50 titers were calculated by fitting a 4-parameter logistic curve.

### Transcriptomic profiling

Metabolic transcriptomic signatures of CD8^+^ T cells were profiled in cells isolated from 4 WT and 4 C5024T mice. Mice were age (2-month-old) and sex matched. Briefly, for each mouse, freshly purified (negative selection) splenic naïve CD8^+^ T cells were seeded at a density of 200,000 cells/well and activated using a 1:1 (bead:cell) ratio with Dynabeads Mouse T-Activator CD3/CD28 for 6 and 96 h at 37°C (5% CO_2_) in the presence of IL-2. 200,000 *ex vivo* purify ed naïve CD8^+^ T cells were collected for baseline (0 h) measurement. Total RNA was extracted using Trizol LS reagent (Invitrogen) and RNA concentrations determined using a NanoDrop ND-1000 UV-Vis Spectrophotometer (ThermoFisher Scientific). The 6 h timepoint for one C5024T sample failed QC (due to low RNA binding) and was excluded from the analyses.

A well-validated 768-gene panel covering 34 annotated pathways involved in five important metabolism themes (mouse nCounter Metabolic Pathways Panel, NanoString Technologies) was used to explore metabolic transcriptome signatures of CD8^+^ T cells *in vitro* after TCR activation. 200 ng of total RNA was incubated at 65 °C for 24 h to hybridize with reporter and capture probes. The RNA complexes were subsequently immobilized and counted on an nCounter analyzer (NanoString Technologies) according to the manufacturer’s instructions. Raw data were normalized and analysed in nSolver 4.0 (NanoString). Data are deposited in GEO (NCBI, https://www.ncbi.nlm.nih.gov/geo/, accession number: GSE182887).

### qPCR

For qPCR of mouse BMDMs, RNA was obtained from Trizol samples are previously described. 500 ng RNA was reverse transcribed using the High-capacity cDNA Reverse Transcription Kits (ThermoFisher Scientific) in a 20 μl reaction. 20 μl cDNA was diluted to 100 μl and 1 μl cDNA was used for the real-time quantitative PCR by using the Platinum SYBR Green qPCR SuperMix-UDG (ThermoFisher Scientific) and PCR machine QuantStudio 6 Flex (ThermoFisher Scientific). Primers (5’-3’) used: *Arg1* F: CTCCAAGCCAAAGTCCTTAGAG; R: AGGAGCTGTCATTAGGGACATC. *Il1b* F: GCAACTGTTCCTGAACTCAACT; R: ATCTTTTGGGGTCCGTCAACT. *MHC-II (IA)* F: GTGGTGCTGATGGTGCTG; R CCATGAACTGGTACACGAAATG. *GAPDH* F: TGAAGCAGGCATCTGAGGG; R: CGAAGGTGGAAGAGTGGGAG. *Beta actin* F: GGCTGTATTCCCCTCCATCG; R: CCAGTTGGTAACAATGCCATGT.

### Statistical analyses

Statistical analyses were carried out in Prism 9 (GraphPad) and results are represented as mean ± SEM. Comparisons between naïve and memory cells from one individual were calculated using paired, two-tailed Student’s t-tests. Unpaired, two-tailed Student’s t-tests were used for group comparisons. Analysis of more than two types of immune cell from one individual were calculated using paired one-way ANOVA with Bonferroni correction. Bonferroni-corrected analyses were used for gene expression data.

Please contact the authors about material or data, available upon request.

## Data Availability

Please contact the authors about material or data, available upon request.

## Acknowledgments

Our gratitude is for the MELAS patients and animals that contributed to the study. The work was funded by Max Planck Institute, Karolinska Institutet (WAF2017, awarded to JR), the Knut & Alice Wallenberg Foundation (KAW 2018.0080, awarded to JR; 2015.0063 awarded to RHo), and the Swedish Research Council (VR2016-02179, awarded to AWr; VR2015-02662, awarded to RHo). Special thanks go to Jonathan Coquet for assistance with mouse vaccinations, Juan Basile for expertise with flow cytometry, and Nils-Göran Larsson, for critical discussion.

## Author information

JZ, GBKH, XCD and JR designed the study and wrote the manuscript with input from co-authors. JZ, XCD and JR carried out wet-lab experiments and analyzed the data in consultation with coauthors. AWe, AWr and ME recruited MELAS patients. CK assisted with animal experiments, along with ST, JH and YL. MAd, MAo and LB assisted with flow cytometry. RF assisted with pyrosequencing. LH, BM and GM provided SARS-CoV-2 antigens and reagents. DJS carried out SARS-CoV-2 neutralization assays. RHa, RHo and MP provided critical reagents and advised on different aspects of the project.

## Resources Table

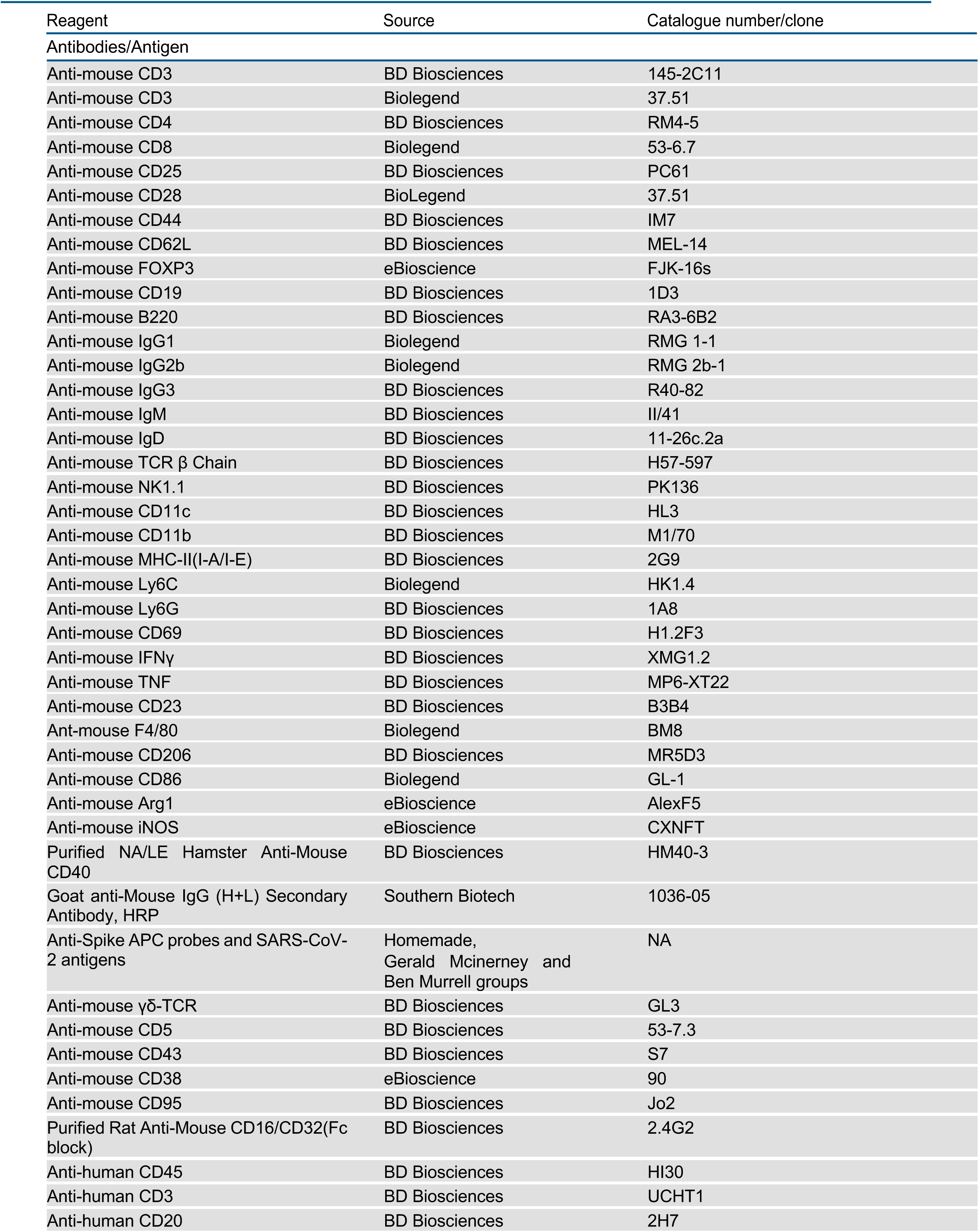
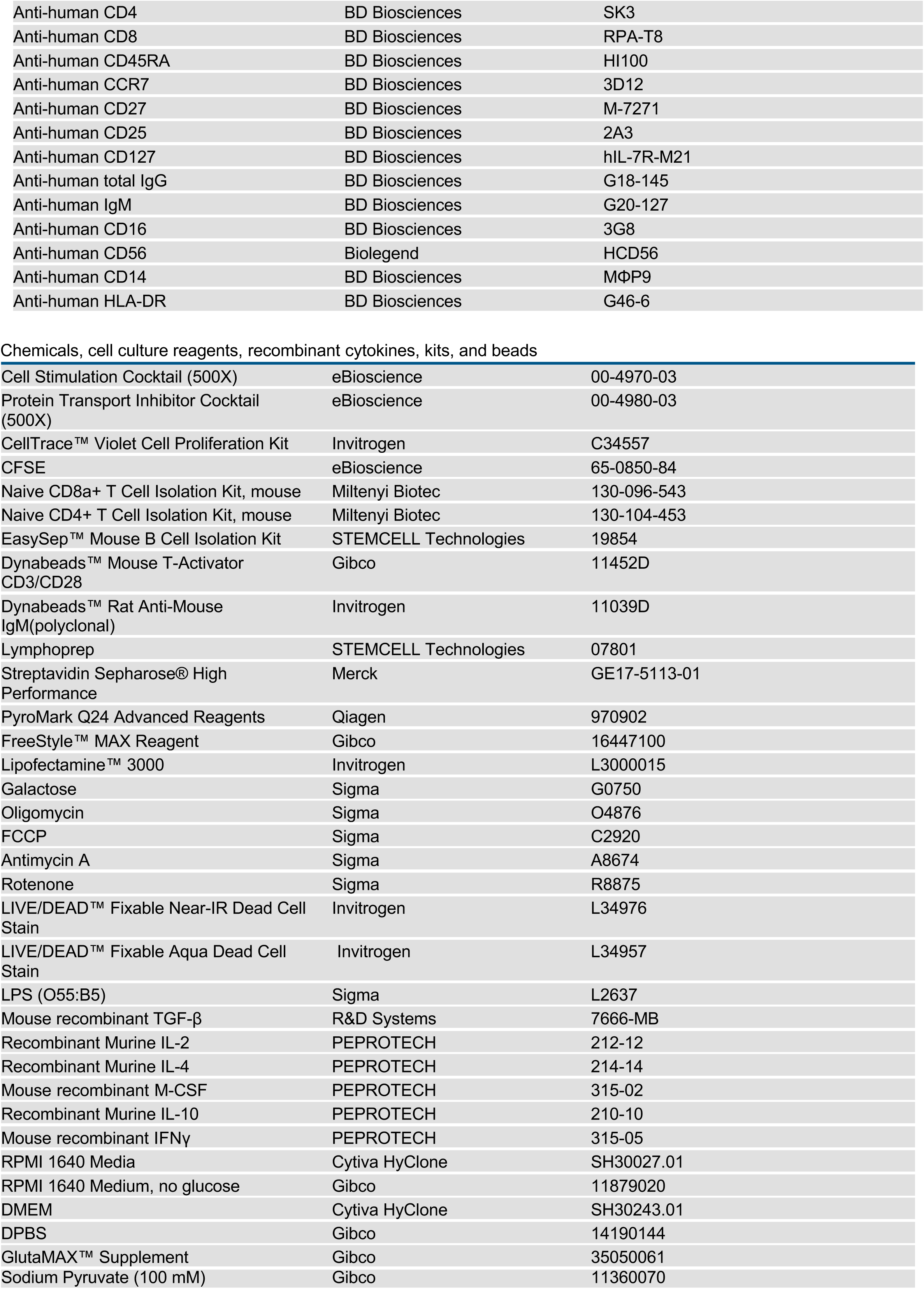
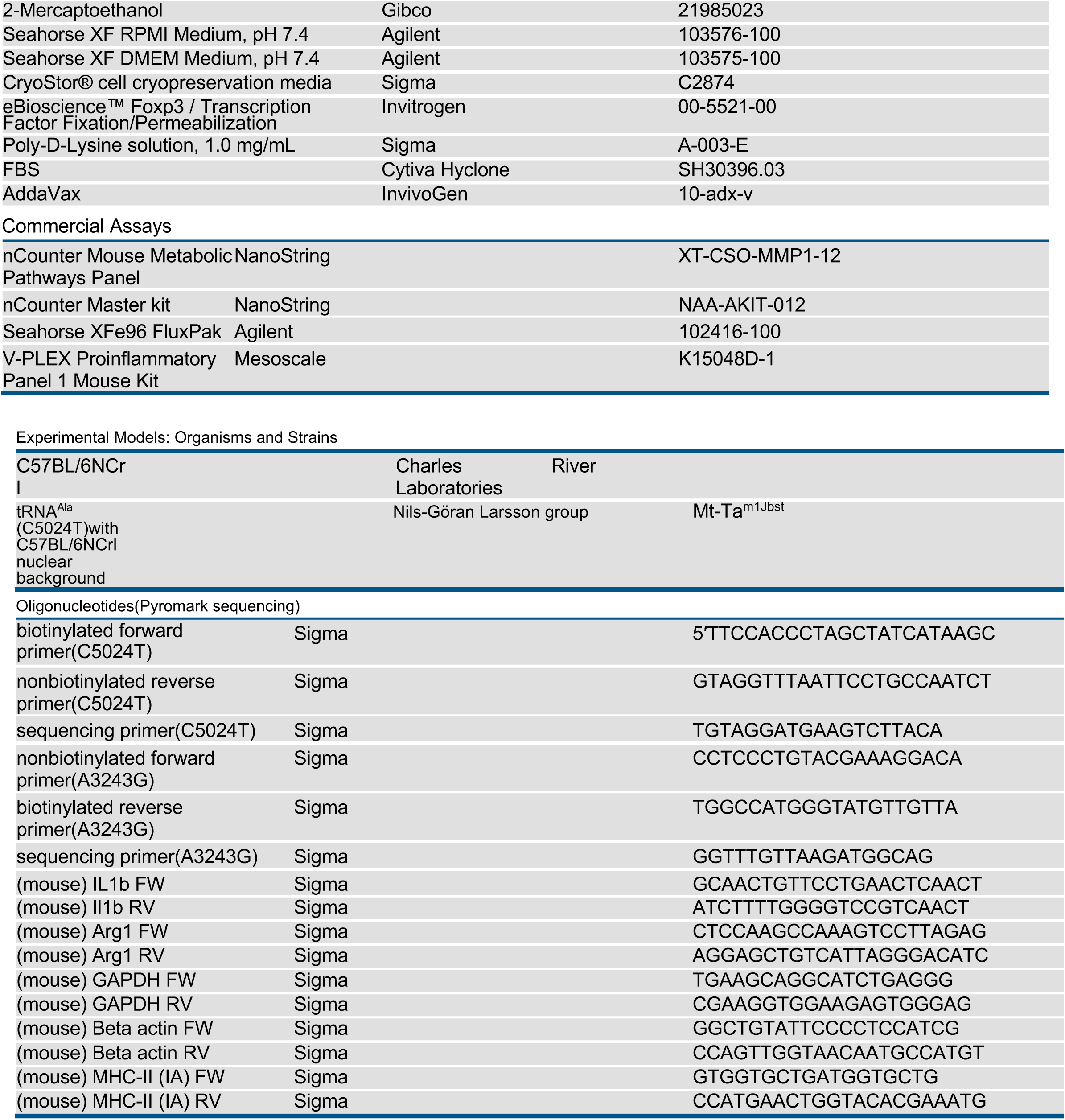

## Graphical Abstract

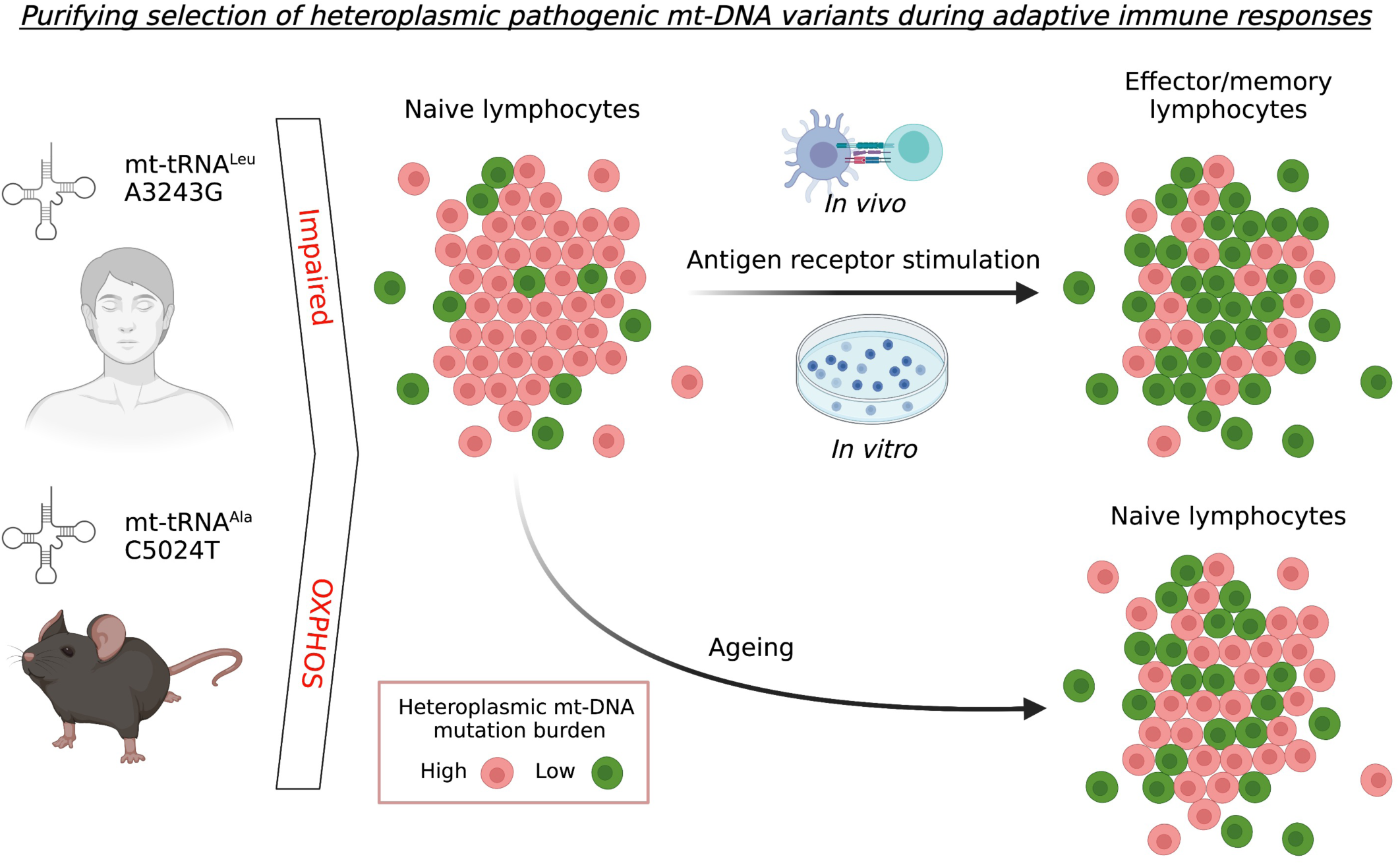

